# SAFETY ASSESSMENT OF LUTEIN AND ZEAXANTHIN SUPPLEMENTATION AND ITS EFFECTS ON BLOOD GLUCOSE LEVELS, KIDNEY FUNCTIONS, LIVER FUNCTIONS AND BONE HEALTH - A RANDOMIZED, DOUBLE-BLIND, PLACEBO-CONTROLLED CLINICAL STUDY

**DOI:** 10.1101/2023.09.22.23295947

**Authors:** C A Anzar, M V Joseph, R Sundaram, G B Vadiraj, C P Prasad, Bineesh Eranimose, Shobhith Jagadeesh

**Affiliations:** Olive Lifesciences Pvt Ltd No.165/5, Near NH-4, Nelamangala, Bangalore – 562123, Karnataka, India; Clinical Research Mallikatta Neuro Centre, Mallikatte, Kadri, Mangalore, Karnataka 575002, India

**Keywords:** Lutein and Zeaxanthin, Type-2 diabetes, Kidney function, Liver health, Bone health

## Abstract

**Objective:** HbA1c, a major marker for the sugar levels in the blood, is the litmus test for people who are on the verge of entering the diabetic zone and for those who are already affected by this disease. Oral hypoglycemic agents are the fine line of treatment in such cases. Nutraceutical and herbal supplements can be utilized as a prophylactic to keep such diseases at bay. Lutein, a carotenoid from the marigold flower, is a very well-known ingredient in the management of eye health. Lutein and zeaxanthin, put together, are commonly known as macular pigments. These pigments help in filtering the blue light, thus protecting the eyes from the harmful effects of the blue light emitted from the screens of electronic gadgets. However, recent studies have demonstrated that these macular pigments have a significant effect on improving cognition and overall brain health. A randomized, double-blind, placebo-controlled clinical study was conducted on lutein and zeaxanthin to determine their effect on cognitive performance. As a safety parameter, HbA1c was also recorded during the study. At the end of the study, the statistics on the data revealed that lutein and zeaxanthin have a positive impact on HbA1c levels. It was observed that the HbA1c of the subjects in the treatment group was significantly lower than that of those in the placebo group, and the values significantly improved during the treatment duration between weeks 1 and 5. As a result, the current study examines how lutein and zeaxanthin affect type 2 diabetes mellitus (T2DM), diabetic kidney disease (DKD), non-alcoholic fatty liver disease (NAFLD), and bone health in healthy individuals between the ages of 35 and 75.

**Methods:** The blood parameters that were measured in thirty individuals who were randomly divided into two groups are the basis for the present study. The trial consisted of two parallel treatment groups and was randomized, double-blind, placebo-controlled clinical research. Through advertising, healthy participants between the ages of 35 and 75 were identified in the community. Following screening, 30 participants were accepted into the trial and randomly assigned using a computer-based randomization methodology into the two model groups, G1 (Group 1-treated) and G2 (Group 2-placebo). The HbA1c level for type 2 diabetes was divided into three groups based on numerical values at various levels: “improved,” “no change,” or “unfavourable.” For instance, if the levels of HbA1c decreased, they were categorized as “improved,” while if they increased, they were categorized as “unfavorable.” The safety profile of the supplement and any potential negative effects on the kidneys are evaluated using renal function testing. The tests help determine whether the supplement is altering kidney function markers such as creatinine, blood urea nitrogen, and serum urea. These factors can show whether the supplement damages the kidneys or affects their ability to function properly. After evaluating the levels of albumin, globulin, bilirubin, SGPT, SGOT, ALP, and GGTP, among various liver function markers, the effects of lutein supplementation on liver function were determined. According to a study by Takeda et al., taking lutein for four weeks significantly increased the amount of cortical bone in the femur and the overall amount of femoral bone mass. This improvement was quantified by dual X-ray absorptiometry and microcomputed tomography (CT) assessments of bone mineral density.

**Results:** A measure of blood sugar levels called estimated average glucose level (eAG), which is generated from HbA1c, showed similar patterns. At visit 1, the treatment group (group 1, Lutein and Zeaxanthin) reported an average eAG of 135.54, whereas the placebo group (group 2, Placebo) exhibited an eAG of 119.98. Before receiving treatment, group-1’s mean eAG was naturally higher than that of group-2. The mean BUN values at visit 1 for the treatment group (group 1, Lutein and Zeaxanthin) were 10.83, while they were 10.13 for the control group (group 2, Placebo). Groups 1 and 2 showed BUN levels of 11.03 and 10.7, respectively, during visit 5. The mean serum urea levels for groups 1 and 2 were 23.2 and 21.69, respectively. The mean values for groups 1 and 2 at visit 5 appeared to be 23.62 and 22.91, respectively, after 5 weeks. When creatinine levels were evaluated at visit 5, they were practically identical to those at visit 1 (1.02 and 0.99 mg/dL for groups 1 and 2, respectively). The mean values for creatinine during visit 1 were 1.03 and 0.985 mg/dL, respectively. At visit 1, the average albumin levels for groups 1 and 2 were 4.32 and 4.61, respectively. The albumin levels in G1 and G2 were 4.62 and 4.77 mg/dL at visit 5, after receiving therapy for 5 weeks. G1 and G2 exhibited total bilirubin levels of 0.609 and 0.547 mg/dL, respectively, during visit 1. For G1 and G2, the respective readings on the visit-5 were 0.633 and 0.662. During visit 1, the total bilirubin levels for G1 and G2 were 0.183 and 0.176 mg/dL, respectively. For G1 and G2, the respective readings during Visit-5 were 0.217 and 0.219 mg/dL. Throughout this clinical trial, there were no severe adverse effects.

**Conclusion:** Clinical investigations have shown that the Lutein and Zeaxanthin is safe for bone, kidney, liver, and diabetes health. It was also noted that the Lutein supplementation helped in managing the HbA1c levels. Thus this study helps in establishing the positive effects of Lutein supplementation in people with impaired blood glucose levels.

## INTRODUCTION

Carotenoids are pigments that are generated from plants and have a number of functions in human biology (1). The xanthophyll carotenoids (XC), lutein (L), and zeaxanthin (Z) are primarily deposited in the human macula. The carotenoid family of nutrients includes lutein and zeaxanthin, which are fat-soluble nutrients. Marigold flowers, egg yolks, corn, and leafy green vegetables with dark green leaves like kale and spinach contain lutein (2). Zeaxanthin can be found in higher concentrations in foods that are yellow or orange, including egg yolks, corn, orange capsicums, tangerines, persimmons, mandarins, and oranges (3, 4). Lutein and zeaxanthin are pigments that are found in the brain, breast, fat tissue, and eyes. Lutein is deposited maximum in our brains, despite the fact that it is not the most prevalent carotenoid in our diet (5). In fact, 66 to 77% of the total carotenoid content in human brain tissue is made up of lutein and zeaxanthin (6). There is growing interest in the neuroprotective benefits of lutein and zeaxanthin since they have been found in the hippocampus, cerebellum, frontal, occipital, and temporal cortices (7–9), and they also have potent antioxidant and anti-inflammatory capacities (10, 11).

One of the most common chronic diseases, diabetes mellitus, is regarded as a major issue in the healthcare system. Diabetes is going to be among the primary causes of morbidity and mortality due to the rising incidence of diabetes mellitus (12). It would appear that a greater emphasis on this issue is essential given that diabetes is more common in women and that it also causes metabolic changes that may result in early menopause and increase hormonal symptoms in women, affecting their health and quality of life (13–15). In order to attain the goals of treatment and manage diabetic complications, these patients should take responsibility for cardiovascular risk factors such as hypertension and dyslipidemia and manage them at the same time as controlling blood sugar (16).

The primary aspect of treatment for DKD (diabetic kidney disease) includes blood pressure regulation, renin-angiotensin system (RAS) inhibitors, and glycemic control. These methods have been shown to successfully lower the risk of disease obtain or progress (17, 18). However, individuals with DKD (diabetic kidney disease) who are intolerant to or insensitive to current pharmacotherapies as well as those with declining renal function but normal-albuminuria have unmet needs (19–21).

Non-alcoholic fatty liver disease (NAFLD) is a spectrum of hepatic illnesses characterized by steatosis without competing etiologies, including alcohol usage, chronic drug use, or viral infections (25). NAFLD is a serious public health concern because it affects around a quarter of the world’s population (26). Given its close connection to insulin resistance and abnormal adipose tissue, NAFLD prevalence has increased along with obesity and diabetes (27). The progressive type of NAFLD, known as non-alcoholic steatohepatitis (NASH), can eventually result in cirrhosis and hepatocellular cancer. As there is currently little evidence to support medication, such as vitamin E and thiazolidinedione, the mainstays of NAFLD treatment are weight loss and physical activity (28). There is not a single drug that has been authorized for the treatment of NAFLD at the moment (30).

Carotenoids, which include lycopene, beta-carotene, lutein, zeaxanthin, and beta-kryptoxanthin, have generated a lot of interest in the field of human nutrition because they function as biological antioxidants and help protect the body from reactive oxygen species (ROS) (31, 32). They also play a protective role in conditions like diabetes and cardiovascular disease (CVD), affecting cellular signalling pathways and influencing the expression of specific genes. Dietary carotenoids are primarily accumulated in the liver, from which they are transported by various lipoproteins for their release into the bloodstream, where they are then deposited and stored in various organs and tissues, including the kidneys, adipose tissue, adrenal glands, testes, skin, and prostate (35).

Lutein is a naturally occurring oxygenated carotenoid with clear anti-inflammatory and antioxidative properties (36). Additionally, it is widely acknowledged that oxidative stress and the inflammatory response are strongly associated with T2DM and DKD (37). However, it is yet unknown whether lutein, T2DM, and DKD in older people are correlated. Although potential therapy targets are obvious, the practitioner is confronted with a bewildering array of research and single drug prescriptions that are difficult to understand, communicate to patients, or incorporate into standard diabetic and DR treatments. Among the 30–40 carotenoid metabolites found in human blood, β-carotene, lycopene, lutein, β-kryptoxanthin, and zeaxanthin share the majority of xanthophylls (such as lutein, zeaxanthin, and astaxanthin), which contain one or more oxygen atoms (38). Research has been done extensively on the chemical characteristics of carotenoids, particularly their strong antioxidant potential. There has recently been an increase in interest in their biological activities in relation to their possible function in the prevention of certain chronic diseases including diabetes and its consequences (39–41).

Carotenoids are strong antioxidants that can eliminate ROS and improve a cell’s capacity to avoid oxidative stress, which is thought to be an emerging treatment approach for people with chronic kidney disease (CKD) (42). It can be divided into two main categories: xanthophylls and carotenes. Xanthophylls, which include lutein, zeaxanthin, and β-kryptoxanthin, are oxygenated terpenes, whereas carotenes, which include β-carotene, α-carotene, and lycopene, as well as other less-studied species, are unoxygenated terpenes (43). The ability of each of these carotenoids to scavenge radicals in three steps—electron transfer, hydrogen abstraction, and addition—has been demonstrated, and they all serve as the major scavengers of ROS (44). A nephroprotective effect of β-carotene has been demonstrated in animal investigations using rats given bromobenzene. There hasn’t been any research done yet to determine whether CKD patient mortality is correlated with carotenoid intake. Therefore, the purpose of this study is to determine whether CKD patient mortality risk can be reduced by carotenoid (lutein and zeaxanthin) intake.

These antioxidants (lutein and zeaxanthin) and their metabolites can build up in the liver and have a favourable impact on hepatocyte metabolism by controlling the cellular oxidative state in certain liver diseases. A major and expanding clinical issue in both industrialized and developing nations, non-alcoholic fatty liver disease (NAFLD) is currently regarded as one of the most prevalent chronic liver disorders in the world.

Lutein (L), a dietary carotenoid that inhibits bone resorption, stimulates bone formation, and increases bone density—most prominently in cortical bone (44). If lutein and zeaxanthin really offer protection against bone loss, as the results from the experimental animal data on young mice suggest, then it would be beneficial to understand the association between lutein and zeaxanthin status and bone health prior to the onset of the degeneration that occurs frequently in aging samples.

There have been an increasing number of clinical trials, preclinical trials, and systematic reviews in the past few years for lutein and zeaxanthin in the treatment of diabetes mellitus (DM), chronic kidney disease (CKD), Non-Alcoholic fatty liver disease (NAFLD), and bone health, but no placebo-controlled trials have been identified anywhere. A randomised placebo controlled clinical study was conducted to evaluate the efficacy of Marigold extract (Lutein and Zeaxanthin on cognitive performance and moods in adults with stroke and parkinsonism. As part of the safety evaluation all the participants in this clinical trial were subjected to undergo regular monitoring of their HbA1c, Liver function and Kidney function tests during their tenure of the clinical trial. The ethical clearance was obtained at Mangala Institutional Ethics Committee vide letter Ref. No. MIEC/V6.1/015 and the trial was registered at CTRI vide registration Number CTRI/2022/06/043208. One of the objectives of this study was to study the safety of Lutein and zeaxanthin supplementation on the blood sugar levels through HbA1c, Liver and Kidney function test.

## MATERIAL AND METHODS

### Trial design

The current study was based on blood parameters taken from 30 people who were randomly divided into two groups. The Lutein and Zeaxanthin lutein supplement was placed in group 1, and the placebo were in group 2. The trial consists of two parallel treatment groups in a randomized, double-blind, placebo-controlled clinical research. Through advertising, healthy people in the community between the ages of 35 and 75 were recruited. A computer-based randomization methodology was used to block-randomize 30 participants into the study’s two treatment groups, G1 and G2, after screening. Age and gender distribution throughout the study arms was equal. Participants were required to meet a number of inclusion criteria, including the capacity to give informed consent, a diagnosis of the required disease, severity, or symptom, and a willingness to adhere by all study guidelines. People had to meet exclusion criteria such as significant renal or hepatic impairment, elective surgery scheduled during the trial or other procedures requiring general anesthesia, or any significant disease or disorder that might danger participants or affect the outcome of the trial. Pregnant, lactating, or intending to become pregnant female trial participants were also excluded. The patient characteristics were represented in Table 1.

**Table 1:**
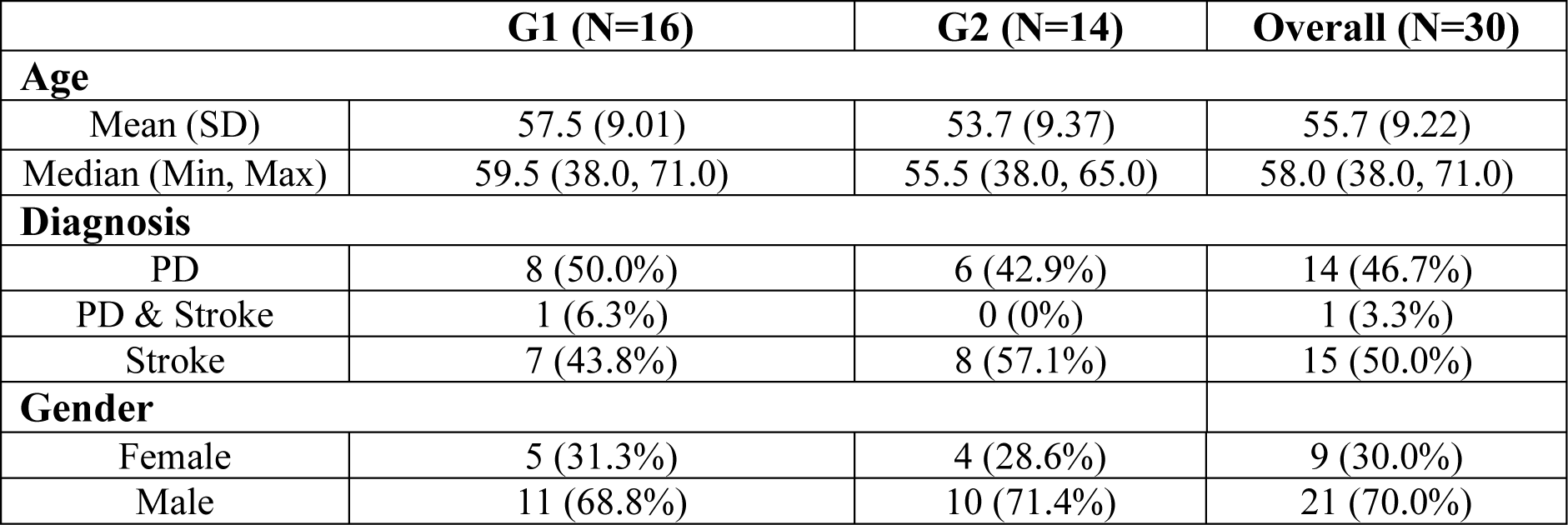
Demographic and Clinical Characteristics of Participants at Study Enrolment.

### Intervention

During the intervention period, participants in the treatment group (G1) received two capsules per day that included 75 mg of lutein and zeaxanthin encapsulated in MCT, standardized to lutein at 10 mg and zeaxanthin at 1 mg. In the placebo group (G2), participants administered capsules that were equivalent to those given to the therapy group. For the course of the study, the capsules were delivered in sealed bottles containing 60 capsules each. The labels on the bottles included an extensive list of details, such as the manufacture date, expiration date, serial number assigned to the trial dose, an explicit declaration that it was only intended for clinical trial purposes and not for commercial use, and a beneficial warning that it was an investigational product. The label also advised against taking a higher dosage, suggested keeping it away from minors and storing it in a cold, dry location, suggested taking the drug with water to prevent swallowing issues, and advised exercising caution if someone suffers any discomfort. The blood parameters were taken at visits 1 and 5, and the tables below illustrate their particulars.

### 1. Lutein and Zeaxanthin Supplementation on Blood Glucose Levels

Low lutein levels were significantly correlated with high glycated hemoglobin (HbA1c) levels, according to a population-based cross-sectional investigation (45). In a different cross-sectional study, 1597 Australian adults were used, and the results revealed an inverse relationship between serum lutein levels and type 2 diabetes (T2DM), as well as a decrease in fasting blood glucose (FBG), fasting insulin, and post-load plasma glucose levels with an increase in lutein quintiles (46). In addition, a recent meta-analysis by Jiang et al. (47) showed that high food intake and circulating lutein levels are linked to a decreased risk of T2DM. Additionally, eight weeks of lutein administration significantly decreased FBG levels in a streptozotocin-induced rat model of T2DM, improved glucose tolerance, and raised the antioxidant enzyme activities in the blood, heart, and kidneys (48). Serum lutein levels have even been suggested as a possible T2DM measure in a recent study (49). The HbA1c level for type 2 diabetes was divided into three groups based on numerical values at various levels: “improved,” “no change,” or “unfavourable.” For instance, if the levels of HbA1c decreased, they were categorized as “improved,” while if they increased, they were categorized as “unfavorable.” If there was no change in HbA1c levels, it was categorized as “no change.” This categorization of the clinical parameter allowed for the use of statistical tests designed for analysing categorical data, suchas Fisher’s exact test, to investigate the relationship between the clinical parameter and treatment. This study also measured random blood sugar levels during the visits.

### 2. Lutein and Zeaxanthin Supplementation On Kidney Function

Lutein could enhance kidney function, according to several studies. In accordance with an investigation published in 2014 (50), the antioxidant and anti-inflammatory properties of lutein have the potential to protect the kidneys from reperfusion from ischemia injury-induced kidney damage. The use of lutein appears to restore kidney function in rats exposed to IRI and considerably lower levels of kidney damage markers such as serum creatinine and blood urea nitrogen. In a recent study (51), serum lutein levels were shown to be substantially reduced in elderly people with type 2 diabetes mellitus (T2DM) and diabetic kidney disease (DKD) compared to healthy controls. Lutein concentrations were found to be a diagnostic sign for T2DM and DKD since they exhibited a negative correlation with a number of metabolic markers. These findings indicate that lutein may act as a preventative measure against the development of T2DM and DKD and may serve as a biomarker for the diagnosis of these conditions. These studies attribute lutein’s anti-inflammatory and antioxidant characteristics to its positive impact on the kidney. Due to the fact that oxidative stress plays a significant role in kidney damage, lutein’s capacity to decrease oxidative stress is suggested as the mechanism underlying its positive benefits. Several kidney function markers, including BUN, serum urea, and creatinine, were assessed in the current study’s two groups at visits one and five, which occurred before and after the various therapies, respectively.

### 1. 3. Lutein and Zeaxanthin Supplementation On Liver Function

One of the possible side effects of pharmacological therapies was drug-induced liver damage (DILI). We evaluated liver function in order to carefully assess the effect of our intervention on participant safety. Blood tests known as liver function tests (LFTs) are performed to assess the health and function of the liver. Clinical trials need LFTs because liver dysfunction or damage can be a significant adverse event linked to numerous drugs and treatments. Monitoring LFTs during a clinical trial can aid in the early detection of potential liver toxicity, enabling timely intervention and reducing the risk to research participants. In our clinical trials, the effects of lutein supplementation on liver function are discussed in the sections that follow. During the clinical trial, a number of tests were measured, including albumin, globulin, bilirubin, SGPT, SGOT, ALP, and GGTP levels.

### 1. 4. Lutein and Zeaxanthin Supplementation On bone health

According to a study by Takeda et al., four weeks of lutein treatment significantly increased the mass of the femoral bones, particularly the cortical bone. This improvement was quantified by dual X-ray absorptiometry and microcomputed tomography (CT) evaluations of bone mineral density (52). Male hip fracture risk has been demonstrated to be negatively linked with dietary intake of the carotenoids α-carotene, β-carotene, and lutein (53). Higher serum carotenoid levels were linked to better bone mineral density in both men and women, according to research by Zhang et al. (54), who observed similar outcomes.

### Statistical analysis

The paired t test in R programming was used for the statistical comparison of the baseline characteristics and outcomes between the 2 groups in visits 1 and 2. Descriptive data analysis was carried out using descriptive statistical methods for all analyses. The complete results of the statistical study have been provided in several tables. A P value less than 0.05 was used to determine whether the results were statistically significant in each case.

## RESULTS AND DISCUSSION RESULTS

### 1 Effect of Lutein and Zeaxanthin Supplementation on Blood Glucose Levels

In the current study, the values of HbA1c were measured in each group during visits 1 and 5. The average HbA1c at visit 1 for the treated group (group 1, Lutein and Zeaxanthin) was 6.35, whereas for the placebo group (group 2, placebo), it was 5.8. The mean HbA1c tended to be on the higher side of group-1 compared to group-2 prior to treatment. Five weeks of treatment with lutein supplements resulted in a statistically significant reduction in HbA1c levels. In group 1, the mean HbA1c levels were reduced to 5.9 as measured at visit 5. However, in group 2 (placebo), the mean HbA1c levels were unchanged. The data are shown in Tables 4 and 5 and Figure 1. At the conclusion of five weeks, the outcomes were categorized as “improved,” “no change,” or “unfavorable.” The patients in G-1, 68.8% exhibited improvement in their HbA1c levels (i.e., their HbA1c levels dropped as a result of the treatment), compared to only 35.7% of those in G2. Unfavorable changes in HbA1c were seen in 57% of G-2 participants. Only 18.8% had an unfavorable change in G-1. This data was subjected to statistical analysis using Fisher’s exact test, and the effect was significant since the p value was found to be 0.048. The data was determined in Table 6.

**Figure1:**
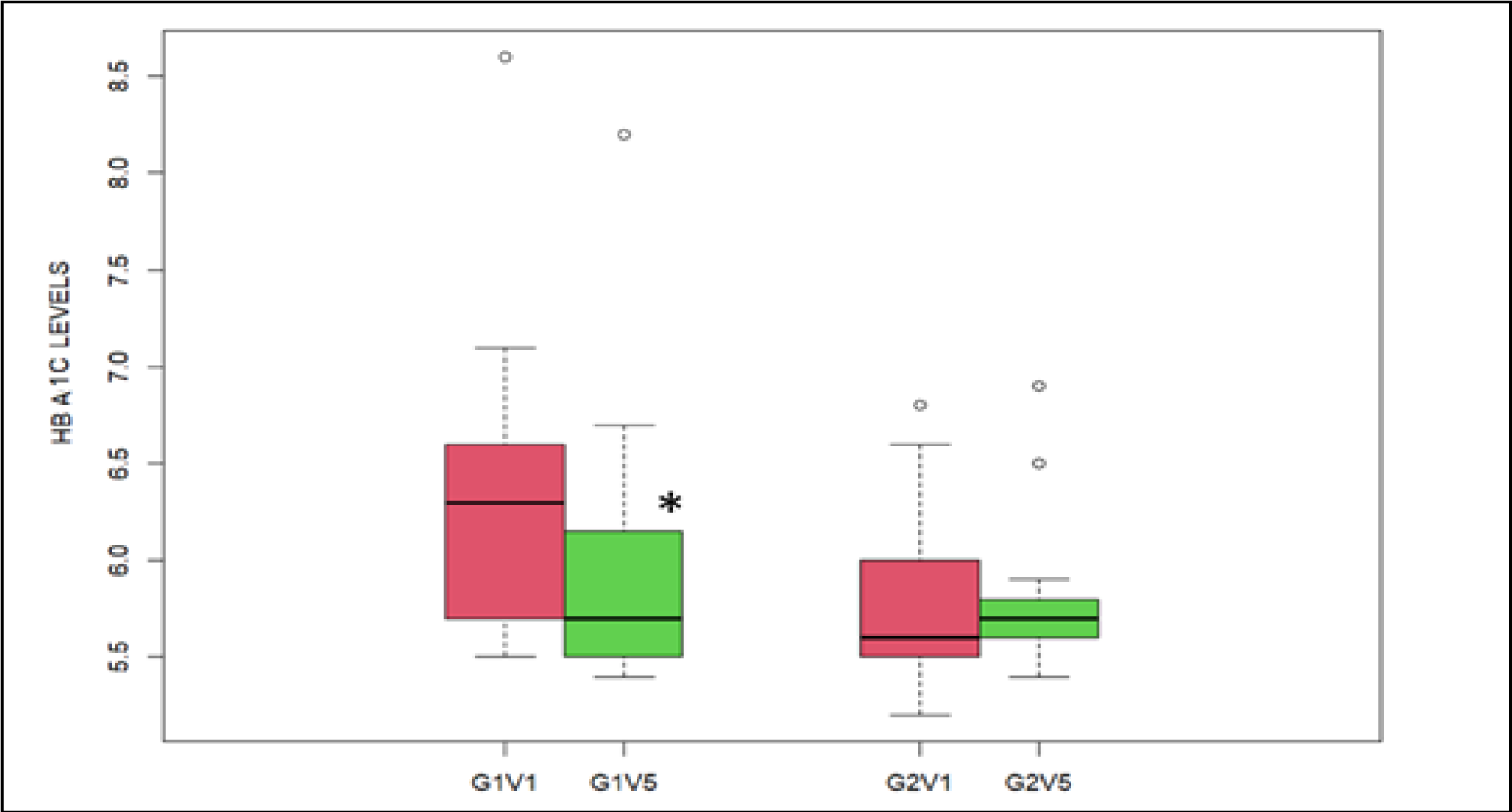
Effect of HbA1c values (mg/dL) during Visit-1 and Visit-5 in G1 (treatment group) and G2 (placebo group)

**Table 2:** Baseline Clinical Parameters of Study Participants (G1, treatment group) and group-2 (G2, placebo group) during visit-1.

|  | **G1 (N=16)** | **G2 (N=14)** | **Overall (N=30)** |
| --- | --- | --- | --- |
| **Age** |  |  |  |
| Mean (SD) | 57.5 (9.01) | 53.7 (9.37) | 55.7 (9.22) |
| Median (Min, Max) | 59.5 (38.0, 71.0) | 55.5 (38.0, 65.0) | 58.0 (38.0, 71.0) |
| **Gender** |  |  |  |
| Female | 5 (31.3%) | 4 (28.6%) | 9 (30.0%) |
| Male | 11 (68.8%) | 10 (71.4%) | 21 (70.0%) |
| **Bun** |  |  |  |
| Mean (SD) | 10.8 (2.88) | 10.1 (1.74) | 10.5 (2.38) |
| Median (Min, Max) | 10.3 (7.40, 18.9) | 9.95 (8.00, 14.5) | 10.1 (7.40, 18.9) |
| Missing | 1 (6.3%) | 0 (0%) | 1 (3.3%) |
| **Urea Serum** |  |  |  |
| Mean (SD) | 23.2 (6.18) | 21.7 (3.72) | 22.5 (5.11) |
| Median (Min, Max) | 22.0 (15.8, 40.5) | 21.3 (17.1, 31.0) | 21.6 (15.8, 40.5) |
| Missing | 1 (6.3%) | 0 (0%) | 1 (3.3%) |
| **Glucose Random** |  |  |  |
| Mean (SD) | 103 (15.6) | 125 (40.6) | 114 (31.9) |
| Median (Min, Max) | 107 (77.0, 132) | 117 (70.0, 228) | 109 (70.0, 228) |
| Missing | 1 (6.3%) | 0 (0%) | 1 (3.3%) |
| **Creatinine** |  |  |  |
| Mean (SD) | 1.03 (0.256) | 0.985 (0.205) | 1.01 (0.230) |
| Median (Min, Max) | 1.11 (0.720, 1.47) | 0.945 (0.720,1.35) | 1.07 (0.720, 1.47) |
| Missing | 1 (6.3%) | 0 (0%) | 1 (3.3%) |
| **HbA1c** |  |  |  |
| Mean (SD) | 6.35 (0.772) | 5.81 (0.459) | 6.11 (0.696) |
| Median (Min, Max) | 6.30 (5.50, 8.60) | 5.60 (5.20, 6.80) | 6.00 (5.20, 8.60) |
| Missing | 0 (0%) | 1 (7.1%) | 1 (3.3%) |
| **eAG** |  |  |  |
| Mean (SD) | 136 (22.2) | 120 (13.2) | 129 (20.0) |
| Median (Min, Max) | 134 (111, 200) | 114 (103, 148) | 126 (103, 200) |
| Missing | 0 (0%) | 1 (7.1%) | 1 (3.3%) |

**Table 3:** Clinical parameters in group-1 (G1, treatment group) and group-2 (G2, placebo group) during visit-5.

|  | **G1 (N=16)** | **G2 (N=14)** | **Overall (N=30)** |
| --- | --- | --- | --- |
| **Age** |  |  |  |
| Mean (SD) | 57.5 (9.01) | 53.7 (9.37) | 55.7 (9.22) |
| Median (Min, Max) | 59.5 (38.0, 71.0) | 55.5 (38.0, 65.0) | 58.0 (38.0, 71.0) |
| **Gender** |  |  |  |
| Female | 5 (31.3%) | 4 (28.6%) | 9 (30.0%) |
| Male | 11 (68.8%) | 10 (71.4%) | 21 (70.0%) |
| **Bun** |  |  |  |
| Mean (SD) | 11.0 (2.69) | 10.7 (1.69) | 10.9 (2.25) |
| Median (Min, Max) | 10.4 (7.60, 16.6) | 10.8 (7.80, 13.5) | 10.4 (7.60, 16.6) |
| **Urea Serum** |  |  |  |
| Mean (SD) | 23.6 (5.76) | 22.9 (3.62) | 23.3 (4.81) |
| Median (Min, Max) | 22.1 (16.3, 35.5) | 23.0 (16.7, 28.9) | 22.1 (16.3, 35.5) |
| **Glucose Random** |  |  |  |
| Mean (SD) | 101 (31.6) | 118 (47.6) | 109 (40.1) |
| Median (Min, Max) | 94.0 (60.0, 177) | 103 (85.0, 237) | 97.5 (60.0, 237) |
| **Creatinine** |  |  |  |
| Mean (SD) | 1.02 (0.236) | 0.989 (0.265) | 1.01 (0.246) |
| Median (Min, Max) | 1.03 (0.650, 1.41) | 0.935 (0.680, 1.40) | 1.01 (0.650, 1.41) |
| **HbA1c** |  |  |  |
| Mean (SD) | 5.94 (0.733) | 5.80 (0.419) | 5.88 (0.602) |
| Median (Min, Max) | 5.70 (5.40, 8.20) | 5.70 (5.40, 6.90) | 5.70 (5.40, 8.20) |
| **eAG** |  |  |  |
| Mean (SD) | 124 (21.0) | 120 (12.0) | 122 (17.2) |
| Median (Min, Max) | 117 (108, 189) | 117 (108, 151) | 117 (108, 189) |

**Table 4:** HbA1c values (mg/dL) during Visit-1 in G1 (treatment group) and G2 (placebo group)

| HbA1c | G1 (N=16) | G2 (N=14) | Overall (N=30) |
| --- | --- | --- | --- |
| Mean (SD) | 6.35 (0.772) | 5.81 (0.459) | 6.11 (0.696) |
| Median (Min, Max) | 6.30 (5.50, 8.60) | 5.60 (5.20,6.80) | 6.00 (5.20, 8.60) |
| Missing | 0 (0%) | 1 (7.1%) | 1 (3.3%) |

**Table 5:** HbA1c values (mg/dL) during Visit-5 in G1 (treatment group) and G2 (placebo group)

| HbA1c | G1 (N=16) | G2 (N=14) | Overall (N=30) |
| --- | --- | --- | --- |
| Mean (SD) | 5.94 (0.733) | 5.80 (0.419) | 5.88 (0.602) |
| Median (Min, Max) | 5.70 (5.40, 8.20) | 5.70 (5.40,6.90) | 5.70 (5.40, 8.20) |

**Table 6:** Effect of Lutein and Zeaxanthin Supplementation on HbA1c after 5 weeks.

| HbA1c Outcome | G1 (N=16) | G2 (N=14) | Overall (N=30) |
| --- | --- | --- | --- |
| Improved | 11 (68.8%) | 5 (35.7%) | 16 (53.3%) |
| No change | 2 (12.5%) | 0 (0%) | 2 (6.7%) |
| Unfavorable | 3 (18.8%) | 8 (57.1%) | 11 (36.7%) |
| Missing | 0 (0%) | 1 (7.1%) | 1 (3.3%) |

G1V1 (HbA1c levels measured in group-1 at visit-1); G1V5 (HbA1c levels measured in group-1 at visit-5); G2V1 (HbA1c levels measured in group-2 at visit-1); G2V5 (HbA1c levels measured in group-2 at visit-5). The central line in the box plots represents the median values, and the rectangular box shows the interquartile range (IQR), which is the middle 50% of the data. The whiskers extend to the minimum and maximum values of the data, except for the outliers that fall beyond the whiskers. Statistical significance was calculated using a paired, two-sided t-test. The * value represents p<0.05 for comparison between HbA1c levels between G1V1 and G1V5.

### 1.1 Effect of estimated Average Glucose (eAG) Levels

Estimated average glucose level (eAG) is a parameter derived from HbA1c; similar trends were observed. The average eAG at visit 1 for the treated group (group 1, Lutein and Zeaxanthin) was 135.54, whereas for the placebo group (group 2, Placebo), it was 119.98. The mean eAG was, of course, on the higher side of group 1 compared to group 2 prior to treatment. Five weeks of treatment with a lutein supplement resulted in a statistically significant reduction in the eAG levels. In group 1, the mean eAG levels decreased to 123.92 as measured at visit 5. However, in group 2 (placebo), the mean eAG levels were unchanged. This is obviously an expected outcome since eAG is a parameter derived from HbA1c. Tables 7 and 8 and the figure below represent the details.

**Table 7:** eAG values (mg/dL) during Visit-1 in G1 (treatment group) and G2 (placebo group)

| eAG | G1 (N=16) | G2 (N=14) | Overall (N=30) |
| --- | --- | --- | --- |
| Mean (SD) | 136 (22.2) | 120 (13.2) | 129 (20.0) |
| Median (Min, Max) | 134 (111, 200) | 114 (103, 148) | 126 (103, 200) |
| Missing | 0 (0%) | 1 (7.1%) | 1 (3.3%) |

**Table 8:** eAG values (mg/dL) during Visit-5 in G1 (treatment group) and G2 (placebo group)

| eAG | G1 (N=16) | G2 (N=14) | Overall (N=30) |
| --- | --- | --- | --- |
| Mean (SD) | 124 (21.0) | 120 (12.0) | 122 (17.2) |
| Median (Min, Max) | 117 (108, 189) | 117 (108, 151) | 117 (108, 189) |

G1V1 (eAG levels measured in group-1 at visit-1); G1V5 (eAG levels measured in group-1 at visit-5); G2V1 (eAG levels measured in group-2 at visit-1); G2V5 (eAG levels measured in group-2 at visit-5). The central line in the box plots represent the median values and the rectangular box shows the interquartile range (IQR), which is the middle 50% of the data. The whiskers extend to the minimum and maximum values of the data, except for the outliers that fall beyond the whiskers. Statistical significance was calculated using paired, two sided t-test. * value represents p<0.05 for comparison between HB1AC levels between G1V1 and G1V5

### 1.2 Effect of Lutein and Zeaxanthin supplementation on Random glucose levels

There were no statistically significant changes for either of the groups between visits 1 and 5. However, it should be noted that random blood sugar levels can only provide a quick and easy snapshot of a person’s blood sugar level at a specific moment in time. However, they are not a reliable method for tracking diabetes or diabetes control over time. This is because random blood sugar levels can be influenced by various factors, such as recent food intake, physical activity, and stress levels. Hence, the results of random blood sugar levels are not generally taken into consideration for validating the effects of any intervention. Tables 9 and 10 and Figure 3 show the proper results.

**Figure2:**
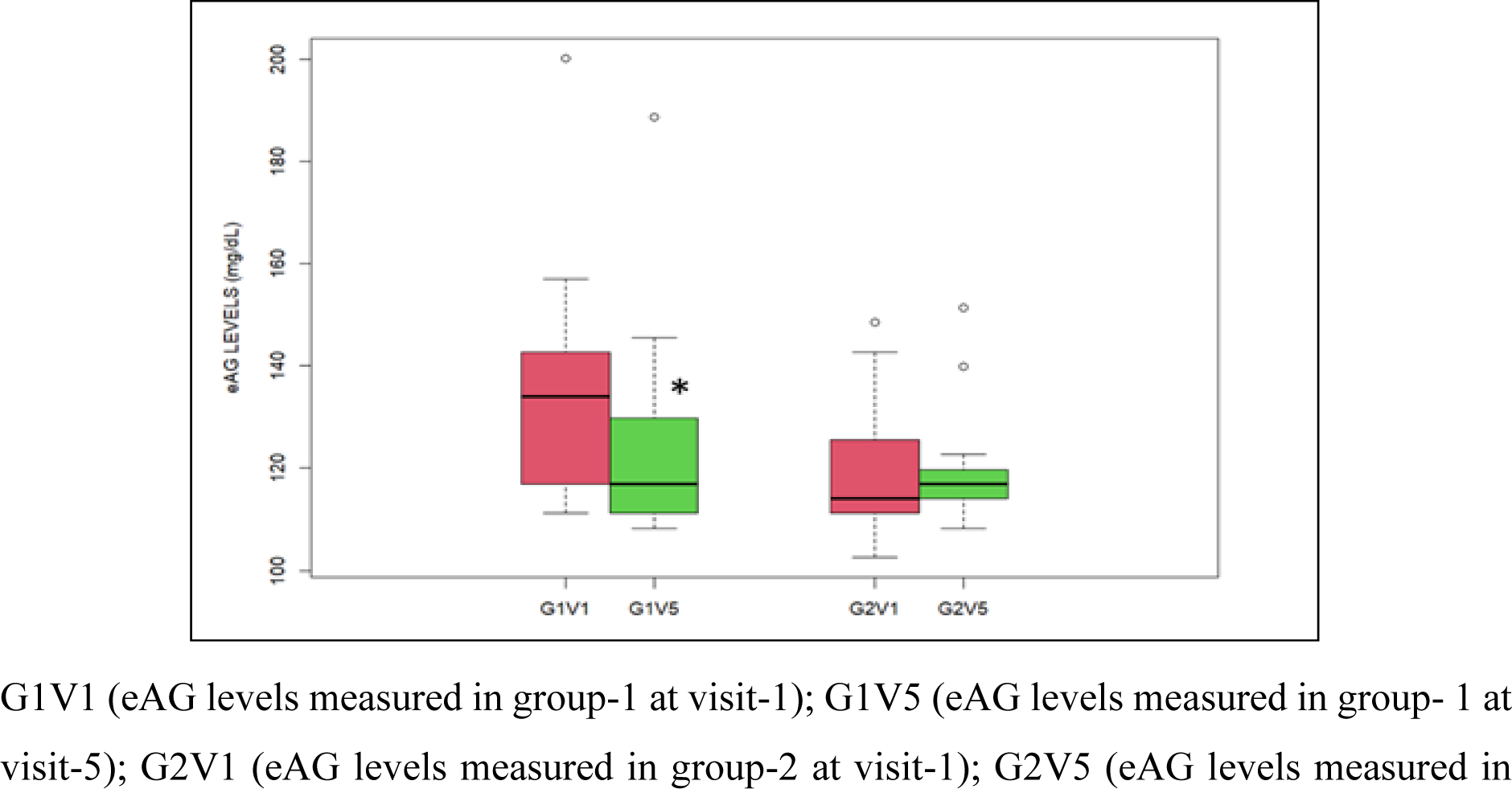
Effect of Lutein and Zeaxanthin Supplementation on Estimated Average Glucose (eAG) levels

**Figure 3:**
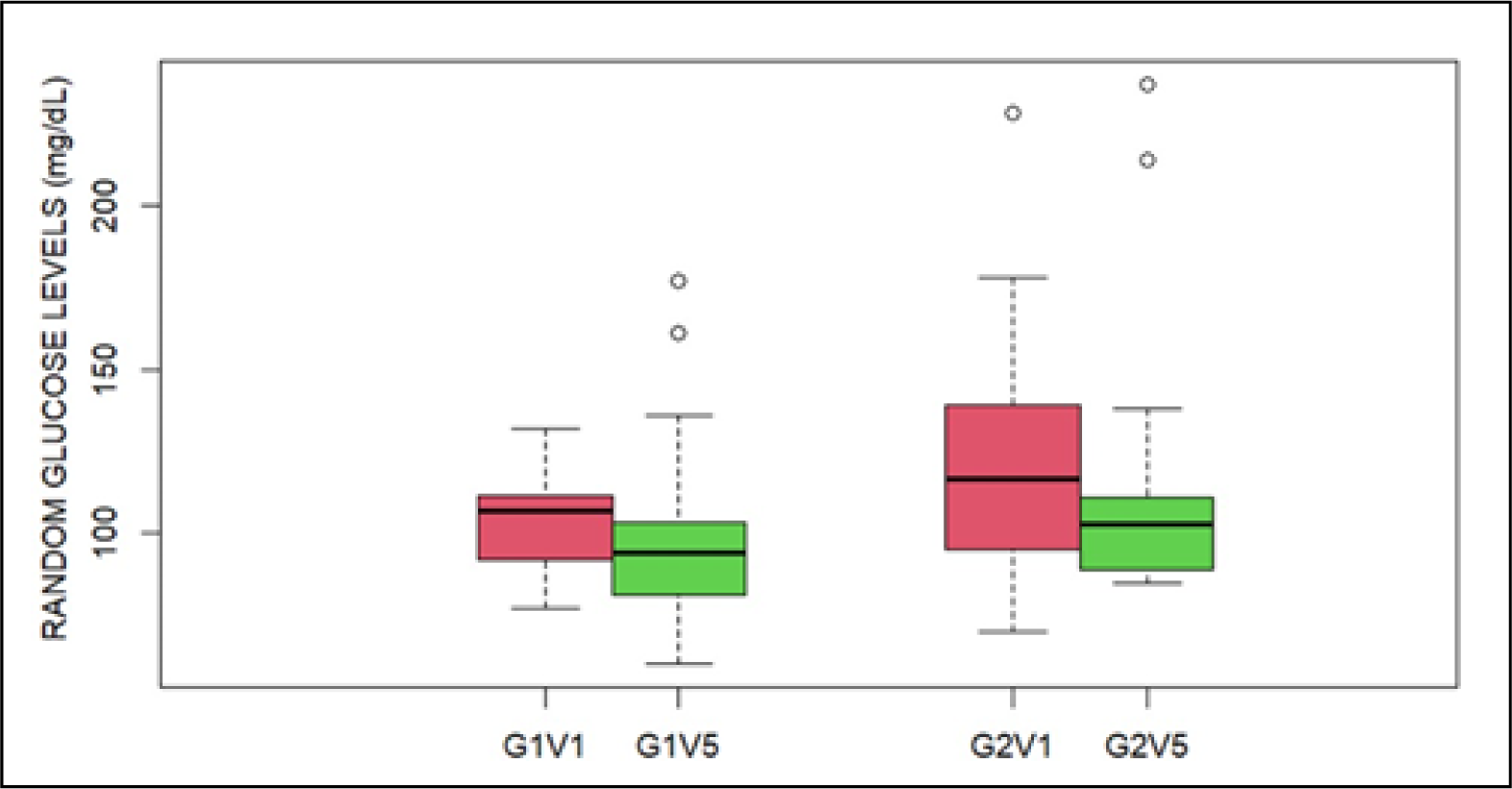
Effect of Lutein and Zeaxanthin supplementation on Random glucose levels

**Table 9:** Random glucose values (mg/dL) during Visit-1 in G1 (treatment group) and G2 (placebo group)

| Random glucose | G1 (N=16) | G2 (N=14) | Overall (N=30) |
| --- | --- | --- | --- |
| Mean (SD) | 103 (15.6) | 125 (40.6) | 114 (31.9) |
| Median (Min, Max) | 107 (77.0, 132) | 117 (70, 228) | 109 (70, 228) |
| Missing | 1 (6.3%) | 0 (0%) | 1 (3.3%) |

**Table 10:** Random glucose values (mg/dL) during Visit-5 in G1 (treatment group) and G2 (placebo group)

| eAG | G1 (N=16) | G2 (N=14) | Overall (N=30) |
| --- | --- | --- | --- |
| Mean (SD) | 101 (31.6) | 118 (47.6) | 109 (40.1) |
| Median (Min, Max) | 94.0 (60, 177) | 103 (85.0, 237) | 97.5 (60.0, 237) |

G1V1 (random blood glucose levels measured in group-1 at visit-1); G1V5 (random blood glucose levels measured in group-1 at visit-5); G2V1 (random blood glucose levels measured in group-2 at visit-1); G2V5 (random blood glucose levels measured in group-2 at visit-5). The central line in the box plots represents the median values, and the rectangular box shows the interquartile range (IQR), which is the middle 50% of the data. The whiskers extend to the minimum and maximum values of the data, except for the outliers that fall beyond the whiskers. Statistical significance was calculated using a paired, two-sided t-test. No significance was observed for corresponding comparisons.

## 2. Effect of Lutein and Zeaxanthin supplementation On Kidney Function

### 2.1 Blood Urea Nitrogen (BUN)

The mean BUN values at visit 1 for group 1 (Lutein and Zeaxanthin) were 10.83, whereas for placebo group 2 (placebo), they were 10.13. During visit 5, the BUN values for group 1 and group 2 were 11.03 and 10.7, as expressed in Tables 11 and 12 and Figure 4, respectively. These changes were not statistically significant as measured by a paired, two-tailed t-test. Blood urea nitrogen (BUN) is a common laboratory test that measures the amount of urea nitrogen in the blood.

**Figure 4:**
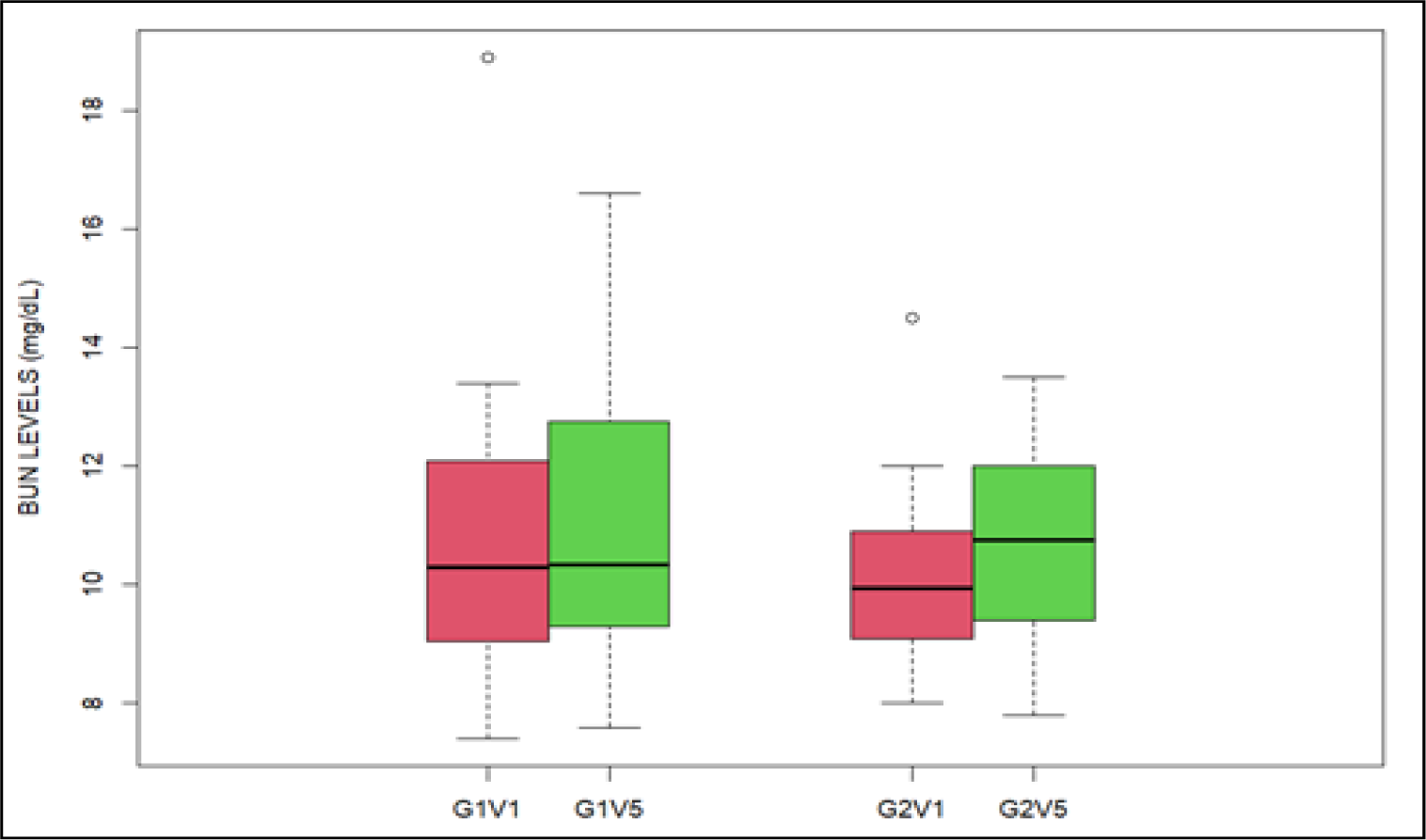
Effect of Lutein and Zeaxanthin supplementation on Blood Urea Nitrogen (BUN) levels

**Table 11:** BUN values (mg/dL) during Visit-1 in G1 (treatment group) and G2 (placebo group)

| BUN | G1 (N=16) | G2 (N=14) | Overall (N=30) |
| --- | --- | --- | --- |
| Mean (SD) | 10.8 (2.88) | 10.1 (1.74) | 10.5 (3.58) |
| Median (Min, Max) | 10.3 (7.40, 18.9) | 9.95 (8.00, 14.5) | 10.1 (7.40, 18.9) |
| Missing | 1 (6.3%) | 0 (0%) | 1 (3.3%) |

**Table 12:** BUN values (mg/dL) during Visit-5 in G1 (treatment group) and G2 (placebo group)

| BUN | G1 (N=16) | G2 (N=14) | Overall (N=30) |
| --- | --- | --- | --- |
| Mean (SD) | 11.0 (2.69) | 10.7 (1.69) | 10.9 (2.25) |
| Median (Min, Max) | 90.4 (7.60, 16.6) | 10.8 (7.80, 13.5) | 10.4 (7.60, 16.6) |

G1V1 (BUN levels measured in group-1 at visit-1); G1V5 (BUN levels measured in group-1 at visit-5); G2V1 (BUN levels measured in group-2 at visit-1); G2V5 (BUN levels measured in group-2 at visit-5). The central line in the box plots represents the median values, and the rectangular box shows the interquartile range (IQR), which is the middle 50% of the data. The whiskers extend to the minimum and maximum values of the data, except for the outliers that fall beyond the whiskers. Statistical significance was calculated using a paired, two-sided t-test. No significance was observed for corresponding comparisons.

### 2.2 Serum Urea levels

The serum urea level is a commonly used laboratory test to assess kidney function and monitor kidney diseases. In the current study, during visit 1, the mean serum urea levels for group 1 and group 2 were 23.2 and 21.69, respectively. At visit 5, after 5 weeks, the mean values for groups 1 and 2 were 23.62 and 22.91, respectively. No significant changes in serum urea levels were observed in either group between the two visits (Tables 13 and 14 and Figure 5).

**Figure 5:**
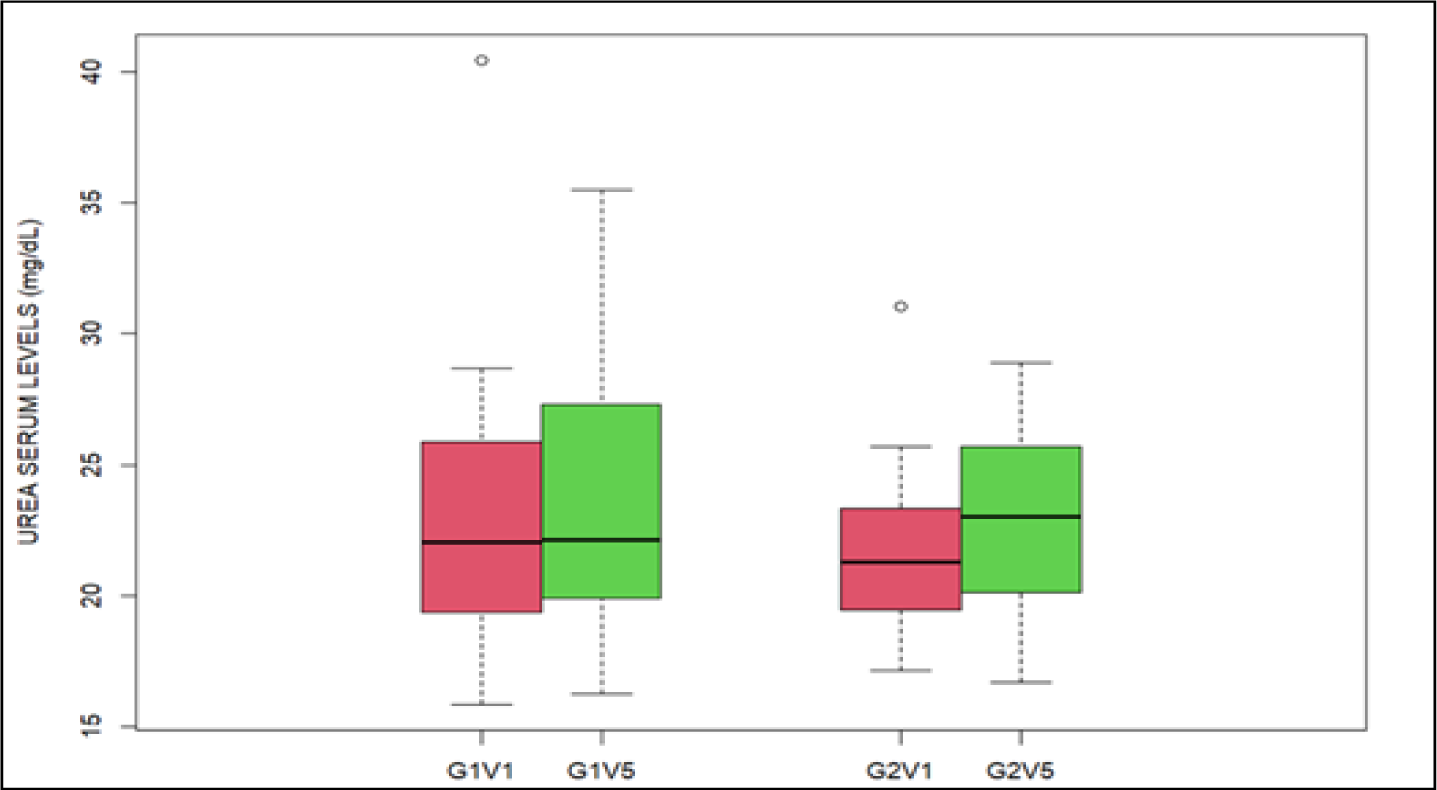
Effect of Lutein and zeaxanthin supplementation on Serum Urea levels

**Table 13:**
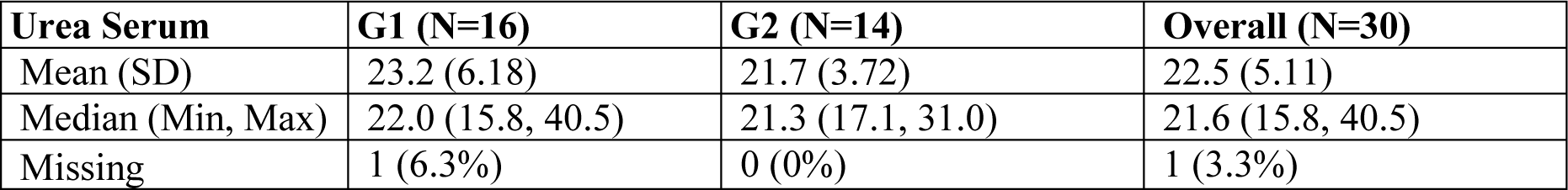
Serum Urea (mg/dL) during Visit-1 in G1 (treatment group) and G2 (placebo group)

**Table 14:** Serum Urea (mg/dL) during Visit-5 in G1 (treatment group) and G2 (placebo group)

| Urea Serum | G1 (N=16) | G2 (N=14) | Overall (N=30) |
| --- | --- | --- | --- |
| Mean (SD) | 23.6 (5.76) | 22.9 (3.62) | 23.3 (4.81) |
| Median (Min, Max) | 22.1 (16.3, 35.5) | 23.0 (16.7, 28.90) | 22.1 (16.3, 35.5) |

G1V1 (serum urea levels measured in group 1 at visit 1); G1V5 (serum urea levels measured in group 1 at visit 5); G2V1 (serum urea levels measured in group 2 at visit 1); G2V5 (serum urea levels measured in group 2 at visit 5). The central line in the box plots represents the median values, and the rectangular box shows the interquartile range (IQR), which is the middle 50% of the data. The whiskers extend to the minimum and maximum values of the data, except for the outliers that fall beyond the whiskers. Statistical significance was calculated using a paired, two-sided t-test. No significance was observed for corresponding comparisons.

### 2.3 Serum creatinine levels

The serum creatinine levels did not show any significant variation in either group during visit 5 in comparison to visit 1, according to the findings of this study. As mentioned in the table below, mean values for creatinine during visit 1 were 1.03 and 0.985 mg/dL, respectively, and the levels remained almost the same when measured at visit 5 (1.02 and 0.99 mg/dL, respectively, for group 1 and group 2) (Tables 15 and 16; Figure 6).

**Figure 6:**
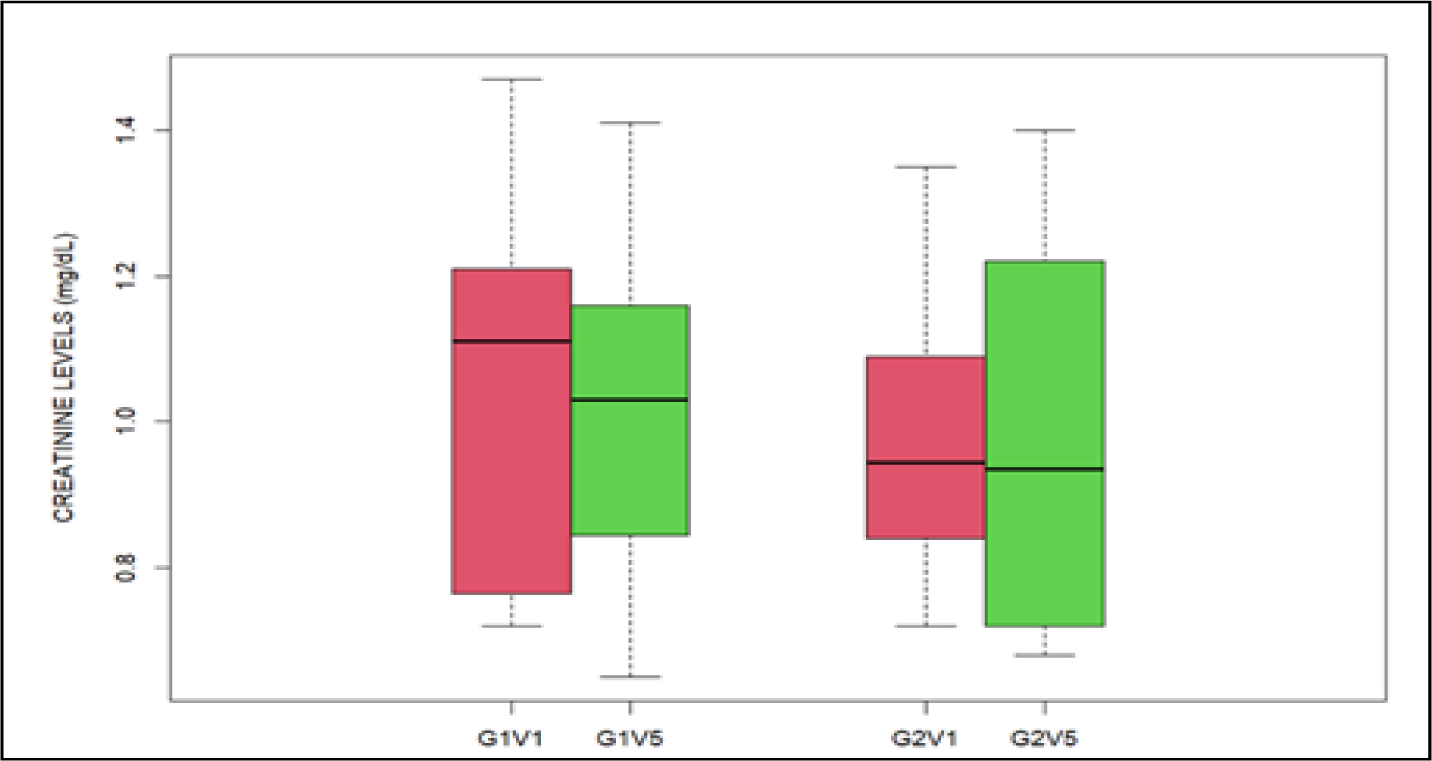
Effect of Lutein and zeaxanthin supplementation on Creatinine levels

**Table 15:** Creatinine levels (mg/dL) during Visit-1 in G1 (treatment group) and G2 (placebo group)

| Creatinine | G1 (N=16) | G2 (N=14) | Overall (N=30) |
| --- | --- | --- | --- |
| Mean (SD) | 1.03 (0.256) | 0.985 (0.205) | 1.01 (0.230) |
| Median (Min, Max) | 1.11 (0.720, 1.47) | 0.945 (0.720, 1.35) | 1.07 (0.720, 1.47) |
| Missing | 1 (6.3%) | 0 (0%) | 1 (3.3%) |

**Table 16:** Creatinine levels (mg/dL) during Visit-5 in G1 (treatment group) and G2 (placebo group)

| Creatinine | G1 (N=16) | G2 (N=14) | Overall (N=30) |
| --- | --- | --- | --- |
| Mean (SD) | 1.02 (0.236) | 0.989 (0.265) | 1.01 (0.246) |
| Median (Min, Max) | 1.03 (0.650, 1.41) | 0.935 (0.680, 1.40) | 1.01 (0.650, 1.41) |

G1V1 (creatinine levels measured in group-1 at visit-1); G1V5 (creatinine levels measured in group-1 at visit-5); G2V1 (creatinine levels measured in group-2 at visit-1); G2V5 (creatinine levels measured in group-2 at visit-5). The central line in the box plots represents the median values, and the rectangular box shows the interquartile range (IQR), which is the middle 50% of the data. The whiskers extend to the minimum and maximum values of the data, except for the outliers that fall beyond the whiskers. Statistical significance was calculated using a paired, two-sided t-test. No significance was observed for corresponding comparisons.

## 3. Effect of Lutein and Zeaxanthin Supplementation On Liver Function

Monitoring LFTs throughout a clinical trial can help identify potential liver toxicity early on, allowing for prompt intervention and minimizing harm to study participants. The following sections describe the effects of lutein supplementation on liver function in our clinical trials. A battery of tests such as levels of albumin, globulin, bilirubin, SGPT, SGOT, ALP, and GGTP were measured during the clinical trial to determine the proper function of the liver. Tables 17 and 18 provide details on the basal levels of these parameters during the study period.

**Table 17:** Liver function test parameters of the participants during visit-1, (G1 – treatment group, G2-Placebo group)

|  | **G1 (N=16)** | **G2 (N=14)** | **Overall(N=30)** |
| --- | --- | --- | --- |
| **Age** |  |  |  |
| Mean (SD) | 57.5 (9.01) | 53.7 (9.37) | 55.7 (9.22) |
| Median [Min, Max] | 59.5 [38.0, 71.0] | 55.5 [38.0, 65.0] | 58.0 [38.0, 71.0] |
| **Total protein** |  |  |  |
| Mean (SD) | 7.24 (0.373) | 7.36 (0.323) | 7.30 (0.348) |
| Median [Min, Max] | 7.09 [6.81, 7.94] | 7.34 [6.64, 7.91] | 7.31 [6.64, 7.94] |
| Missing | 1(6.3%) | 0 (0%) | 1 (3.3%) |
| **Albumin** |  |  |  |
| Mean (SD) | 4.32 (0.251) | 4.61 (0.245) | 4.46 (0.286) |
| Median [Min, Max] | 4.25 [4.03, 4.91] | 4.64 [4.02, 5.10] | 4.45 [4.02, 5.10] |
| Missing | 1 (6.3%) | 0 (0%) | 1 (3.3%) |
| **Globulin** |  |  |  |
| Mean (SD) | 2.93 (0.322) | 2.74 (0.281) | 2.84 (0.312) |
| Median [Min, Max] | 2.86 [2.57, 3.87] | 2.72 [2.27, 3.31] | 2.78 [2.27, 3.87] |
| Missing | 1(6.3%) | 0 (0%) | 1 (3.3%) |
| **Albumin-Globulin ratio** |  |  |  |
| Mean (SD) | 1.49 (0.174) | 1.70 (0.229) | 1.59 (0.225) |
| Median [Min, Max] | 1.51 [1.05, 1.76] | 1.68 [1.39, 2.25] | 1.54 [1.05, 2.25] |
| Missing | 1 (6.3%) | 0 (0%) | 1 (3.3%) |
| **Total Bilirubin** |  |  |  |
| Mean (SD) | 0.609 (0.359) | 0.547 (0.315) | 0.580 (0.335) |
| Median [Min, Max] | 0.480 [0.240, 1.59] | 0.470 [0.240, 1.42] | 0.470 [0.240, 1.59] |
| **Direct Bilirubin** |  |  |  |
| Mean (SD) | 0.183 (0.0814) | 0.176 (0.0803) | 0.180 (0.0795) |
| Median [Min, Max] | 0.160 [0.100, 0.410] | 0.145 [0.0900, 0.360] | 0.155 [0.0900, 0.410] |
| **Indirect Bilirubin** |  |  |  |
| Mean (SD) | 0.426 (0.279) | 0.371 (0.241) | 0.400 (0.259) |
| Median [Min, Max] | 0.315 [0.130, 1.18] | 0.310 [0.150, 1.06] | 0.310 [0.130, 1.18] |
| **SGPT** |  |  |  |
| Mean (SD) | 20.4 (7.36) | 31.4 (15.2) | 25.5 (12.8) |
| Median [Min, Max] | 18.0 [11.0, 38.8] | 30.5 [8.70, 62.0] | 23.0 [8.70, 62.0] |
| **SGOT** |  |  |  |
| Mean (SD) | 21.5 (5.79) | 24.3 (5.88) | 22.8 (5.90) |
| Median [Min, Max] | 21.5 [14.0, 35.0] | 23.5 [16.2, 41.0] | 22.0 [14.0, 41.0] |
| **ALP** |  |  |  |
| Mean (SD) | 77.9 (21.1) | 70.2 (16.3) | 74.2 (19.0) |
| Median [Min, Max] | 74.0 [49.0, 123] | 65.0 [46.0, 108] | 67.0 [46.0, 123] |
| Missing | 1 (6.3%) | 0 (0%) | 1 (3.3%) |
| **GGTP** |  |  |  |
| Mean (SD) | 25.9 (18.8) | 33.3 (22.0) | 29.5 (20.4) |
| Median [Min, Max] | 22.0 [11.0, 90.0] | 26.5 [11.0, 83.0] | 25.0 [11.0, 90.0] |
| Missing | 1 (6.3%) | 0 (0%) | 1 (3.3%) |

**Table 18:** Liver function test parameters of the participants during visit-5, (G1 – treatment group, G2-Placebo group)

|  | **G1 (N=16)** | **G2 (N=14)** | **Overall (N=30)** |
| --- | --- | --- | --- |
| **Age** |  |  |  |
| Mean (SD) | 57.5 (9.01) | 53.7 (9.37) | 55.7 (9.22) |
| Median [Min, Max] | 59.5 [38.0, 71.0] | 55.5 [38.0, 65.0] | 58.0 [38.0, 71.0] |
| **Total protein** |  |  |  |
| Mean (SD) | 7.54 (0.400) | 7.73 (0.343) | 7.62 (0.381) |
| Median [Min, Max] | 7.47 [6.97, 8.25] | 7.59 [7.33, 8.32] | 7.59 [6.97, 8.32] |
| Missing | 0 (0%) | 1 (7.1%) | 1 (3.3%) |
| **Albumin** |  |  |  |
| Mean (SD) | 4.62 (0.281) | 4.77 (0.200) | 4.69 (0.254) |
| Median [Min, Max] | 4.56 [4.26, 5.14] | 4.74 [4.45, 5.15] | 4.69 [4.26, 5.15] |
| Missing | 0 (0%) | 1 (7.1%) | 1 (3.3%) |
| **Globulin** |  |  |  |
| Mean (SD) | 2.92 (0.401) | 2.96 (0.322) | 2.94 (0.362) |
| Median [Min, Max] | 2.79 [2.21, 3.90] | 3.03 [2.47, 3.48] | 2.80 [2.21, 3.90] |
| Missing | 0 (0%) | 1 (7.1%) | 1 (3.3%) |
| **Albumin-Globulin ratio** |  |  |  |
| Mean (SD) | 1.62 (0.260) | 1.63 (0.206) | 1.62 (0.233) |
| Median [Min, Max] | 1.61 [1.12, 2.15] | 1.66 [1.35, 2.01] | 1.65 [1.12, 2.15] |
| Missing | 0 (0%) | 1 (7.1%) | 1 (3.3%) |
| **Total Bilirubin** |  |  |  |
| Mean (SD) | 0.633 (0.426) | 0.662 (0.427) | 0.646 (0.419) |
| Median [Min, Max] | 0.505 [0.300, 2.04] | 0.420 [0.230, 1.45] | 0.500 [0.230, 2.04] |
| Missing | 0 (0%) | 1 (7.1%) | 1 (3.3%) |
| **Direct Bilirubin** |  |  |  |
| Mean (SD) | 0.217 (0.0993) | 0.219 (0.0993) | 0.218 (0.0975) |
| Median [Min, Max] | 0.195 [0.110, 0.500] | 0.180 [0.100, 0.420] | 0.190 [0.100, 0.500] |
| Missing | 0 (0%) | 1 (7.1%) | 1 (3.3%) |
| **Indirect Bilirubin** |  |  |  |
| Mean (SD) | 0.416 (0.333) | 0.443 (0.333) | 0.428 (0.327) |
| Median [Min, Max] | 0.325 [0.190, 1.54] | 0.280 [0.100, 1.06] | 0.310 [0.100, 1.54] |
| Missing | 0 (0%) | 1 (7.1%) | 1 (3.3%) |
| **SGPT** |  |  |  |
| Mean (SD) | 28.2 (14.2) | 30.2 (13.7) | 29.1 (13.7) |
| Median [Min, Max] | 27.0 [14.0, 61.0] | 27.0 [10.1, 55.0] | 27.0 [10.1, 61.0] |
| Missing | 0 (0%) | 1 (7.1%) | 1 (3.3%) |
| **SGOT** |  |  |  |
| Mean (SD) | 26.1 (13.9) | 27.0 (8.70) | 26.5 (11.6) |
| Median [Min, Max] | 23.3 [13.0, 72.0] | 26.0 [17.4, 51.0] | 25.0 [13.0, 72.0] |
| Missing | 0 (0%) | 1 (7.1%) | 1 (3.3%) |
| **ALP** |  |  |  |
| Mean (SD) | 87.2 (25.7) | 68.4 (18.7) | 78.8 (24.4) |
| Median [Min, Max] | 80.0 [48.0, 133] | 67.0 [43.0, 115] | 75.0 [43.0, 133] |
| Missing | 0 (0%) | 1 (7.1%) | 1 (3.3%) |
| **GGTP** |  |  |  |
| Mean (SD) | 29.6 (12.5) | 34.8 (23.5) | 31.9 (18.1) |
| Median [Min, Max] | 25.5 [11.0, 49.0] | 28.0 [10.0, 79.0] | 27.0 [10.0, 79.0] |
| Missing | 0 (0%) | 1 (7.1%) | 1 (3.3%) |

### 3.1 Total protein

Abnormally low levels of total protein may indicate a problem with protein digestion or absorption or a liver or kidney disorder. In our clinical trial, the initial and end-of-study levels of total protein are as represented in Tables 19 and 20.

**Table 19:** Total protein levels (mg/dL) during Visit-1 in group-1 (G1, treatment group) and group-2 (G2, placebo group)

| Total Protein | G1 (N=16) | G2 (N=14) | Overall (N=30) |
| --- | --- | --- | --- |
| Mean (SD) | 7.24 (0.373) | 7.36 (0.323) | 7.30 (0.348) |
| Median (Min, Max) | 7.09 (6.81, 7.94) | 7.34 (6.64, 7.91) | 7.31 (6.64, 7.94) |
| Missing | 1 (6.3%) | 0 (0%) | 1 (3.3%) |

**Table 20:** Total protein levels (mg/dL) during Visit-5 in group-1 (G1, treatment group) and group-2 (G2,placebo group)

| Total Protein | G1 (N=16) | G2 (N=14) | Overall (N=30) |
| --- | --- | --- | --- |
| Mean (SD) | 7.54 (0.400) | 7.73 (0.343) | 7.62 (0.381) |
| Median (Min, Max) | 7.47 (6.97, 8.25) | 7.59 (7.33, 8.32) | 7.59 (6.97, 8.32) |
| Missing | 0 (0%) | 1 (7.1%) | 1 (3.3%) |

G1V1 (total protein levels measured in group-1 at visit-1); G1V5 (total protein levels measured in group-1 at visit-5); G2V1 (total protein levels measured in group-2 at visit-1); G2V5 (total protein levels measured in group-2 at visit-5). The central line in the box plots represents the median values, and the rectangular box shows the interquartile range (IQR), which is the middle 50% of the data. The whiskers extend to the minimum and maximum values of the data, except for the outliers that fall beyond the whiskers. Statistical significance was calculated using a paired, two-sided t-test. *** indicates p<0.001, ## indicates p<0.01

### 3.2 Serum Albumin and globulin levels

As indicated in the two tables below, the average albumin levels in groups 1 and 2 at visit 1 were 4.32 and 4.61, respectively. Following treatment for 5 weeks, at visit 5, the albumin levels in G1 and G2 were 4.62 and 4.77 mg/Dl, respectively (Tables 21 and 22 and Figure 8). Serum globulin levels were also measured during the clinical study, and the levels are as indicated below for G1 and G2 during visits 1 and 5 (Tables 23 and 24 and Figure 9).

**Figure7:**
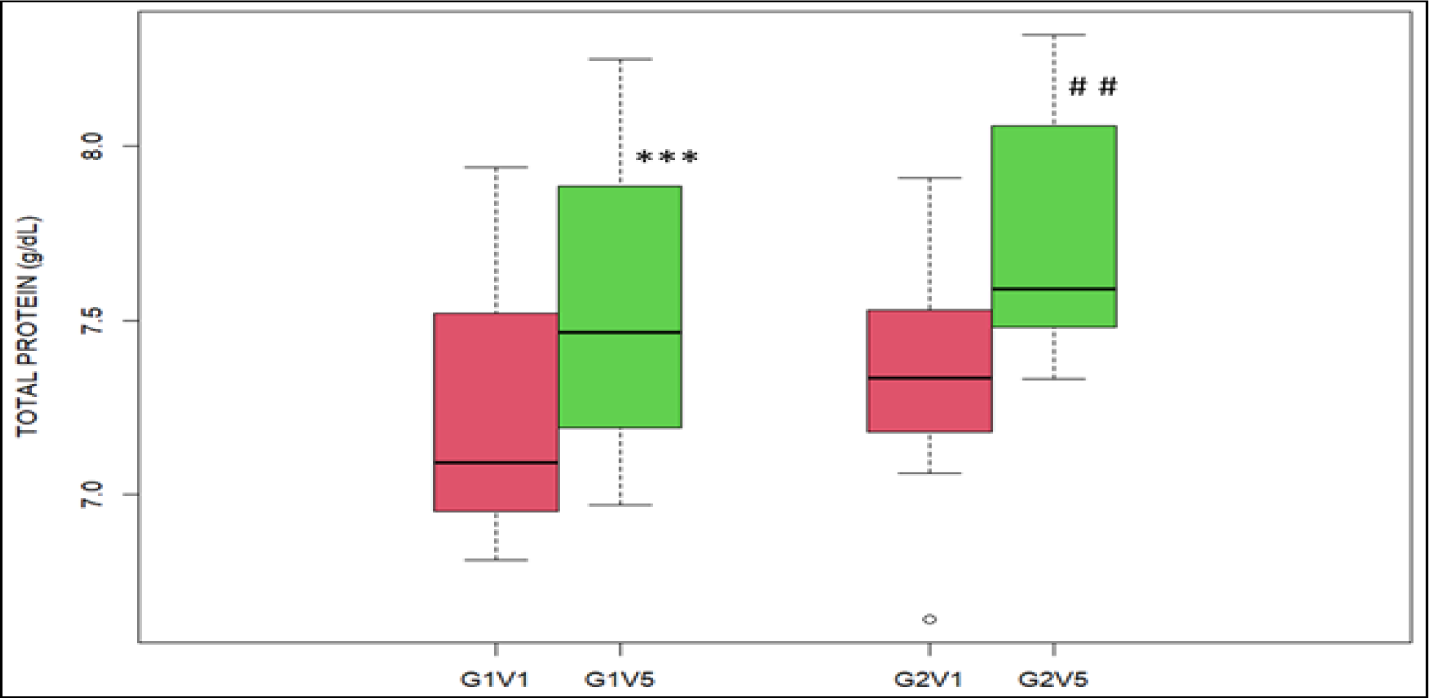
Effect of Lutein and Zeaxanthin supplementation on Total protein levels

**Figure 8:**
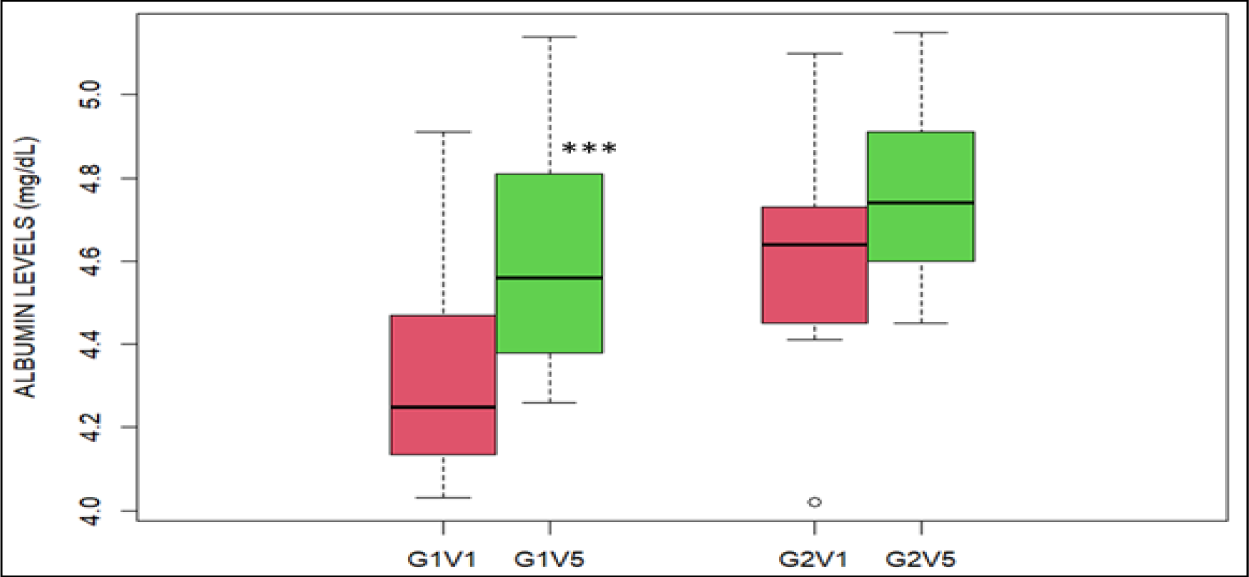
Effect of Lutein supplementation on Albumin levels

**Figure 9:**
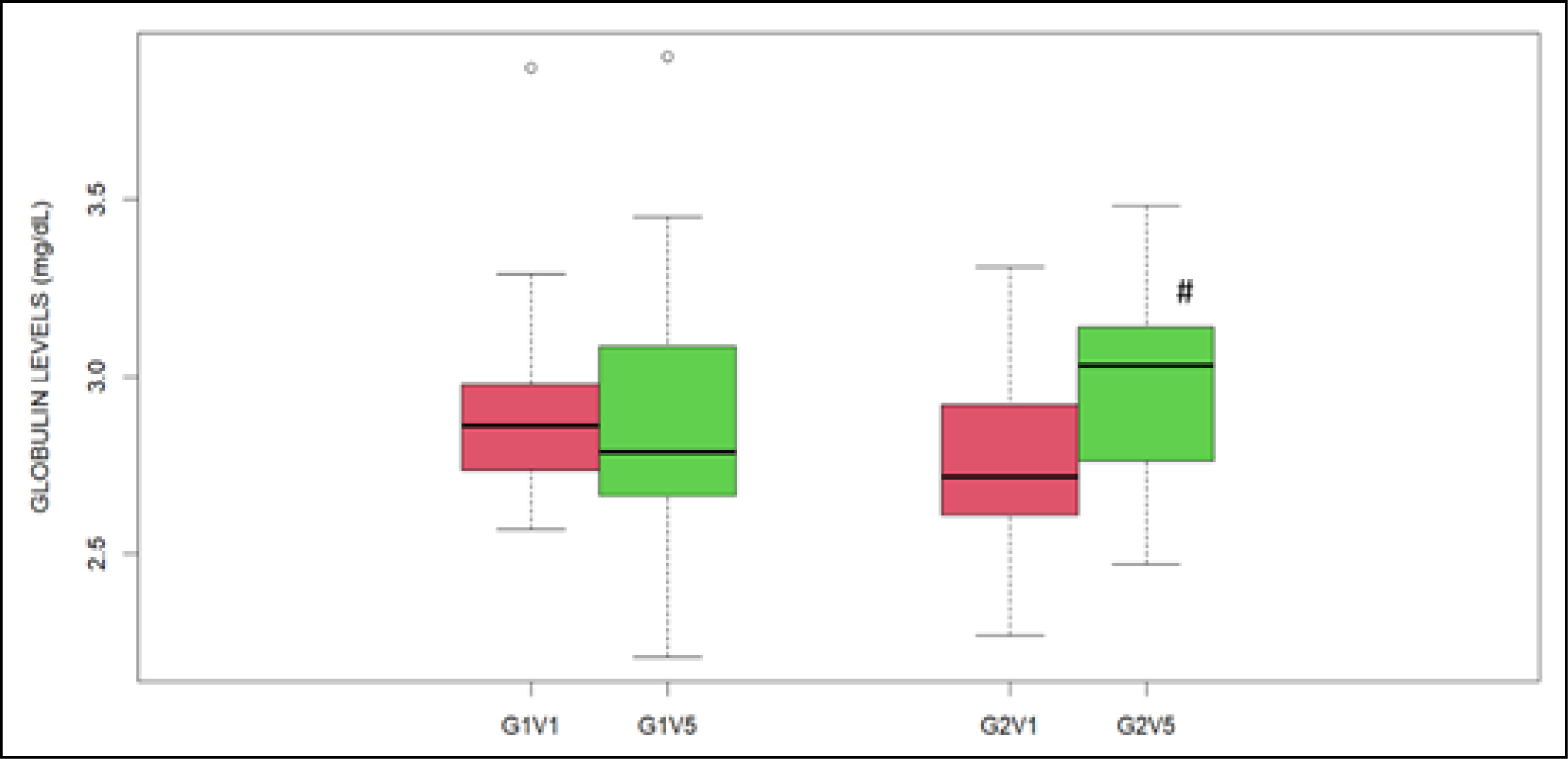
Effect of Lutein and Zeaxanthin supplementation on Globulin levels

**Table 21:**
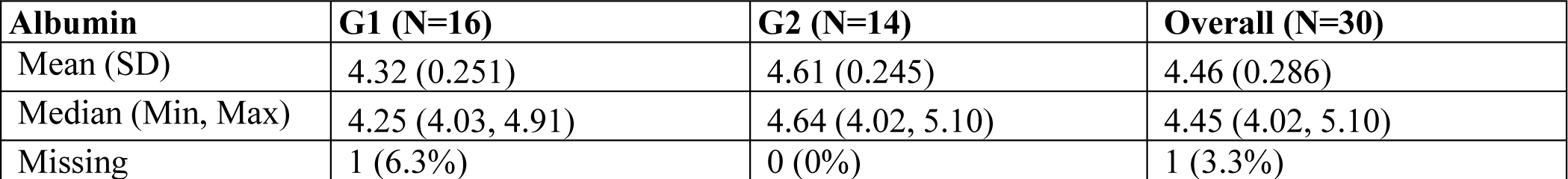
Serum albumin levels (mg/dL) during Visit-1 in group-1 (G1, treatment group) and group-2 (G2, placebo group.

**Table 22:**
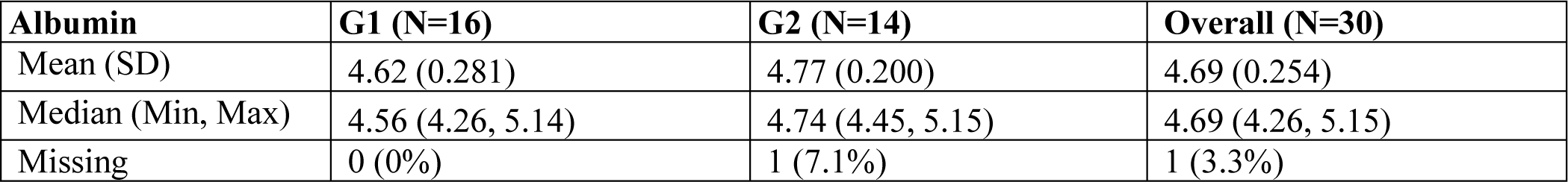
Serum albumin levels (mg/dL) during Visit-5 in group-1 (G1, treatment group) and group-2 (G2, placebo group.

**Table 23:**
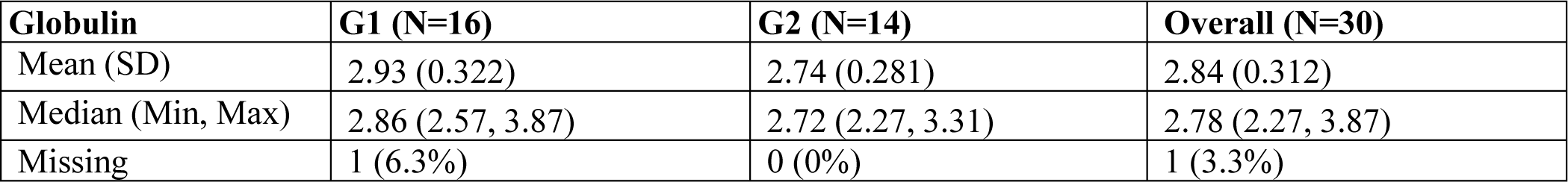
Serum globulin levels (mg/dL) during Visit-1 in group-1 (G1, treatment group) and group-2 (G2, placebo group)

**Table 24:** Serum globulin levels (mg/dL) during Visit-5 in group-1 (G1, treatment group) and group-2 (G2, placebo group)

| Globulin | G1 (N=16) | G2 (N=14) | Overall (N=30) |
| --- | --- | --- | --- |
| Mean (SD) | 2.92 (0.401) | 2.96 (0.322) | 2.94 (0.362) |
| Median (Min, Max) | 2.79 (2.21, 3.90) | 3.03 (2.47, 3.48) | 2.80 (2.21, 3.90) |
| Missing | 0 (0%) | 1 (7.1%) | 1 (3.3%) |

G1V1 (Albumin levels measured in group 1 at visit 1); G1V5 (Albumin levels measured in group 1 at visit 5); G2V1 (Albumin levels measured in group 2 at visit 1); G2V5 (Albumin levels measured in group 2 at visit 5). The central line in the box plots represents the median values, and the rectangular box shows the interquartile range (IQR), which is the middle 50% of the data. The whiskers extend to the minimum and maximum values of the data, except for the outliers that fall beyond the whiskers. Statistical significance was calculated using a paired, two-sided t-test. *** indicates p<0.001

G1V1 (globulin levels measured in group-1 at visit-1); G1V5 (globulin levels measured in group-1 at visit-5); G2V1 (globulin levels measured in group-2 at visit-1); G2V5 (globulin levels measured in group-2 at visit-5). The central line in the box plots represents the median values, and the rectangular box shows the interquartile range (IQR), which is the middle 50% of the data. The whiskers extend to the minimum and maximum values of the data, except for the outliers that fall beyond the whiskers. Statistical significance was calculated using a paired, two-sided t-test. # indicates p<0.05

### 3.3 Albumin Globulin ratio

Tables 25 and 26 and Figure 10 indicate the values of the albumin-to-globulin ratio (AG ratio) for two groups during visits 1 and 5.

**Figure 10:**
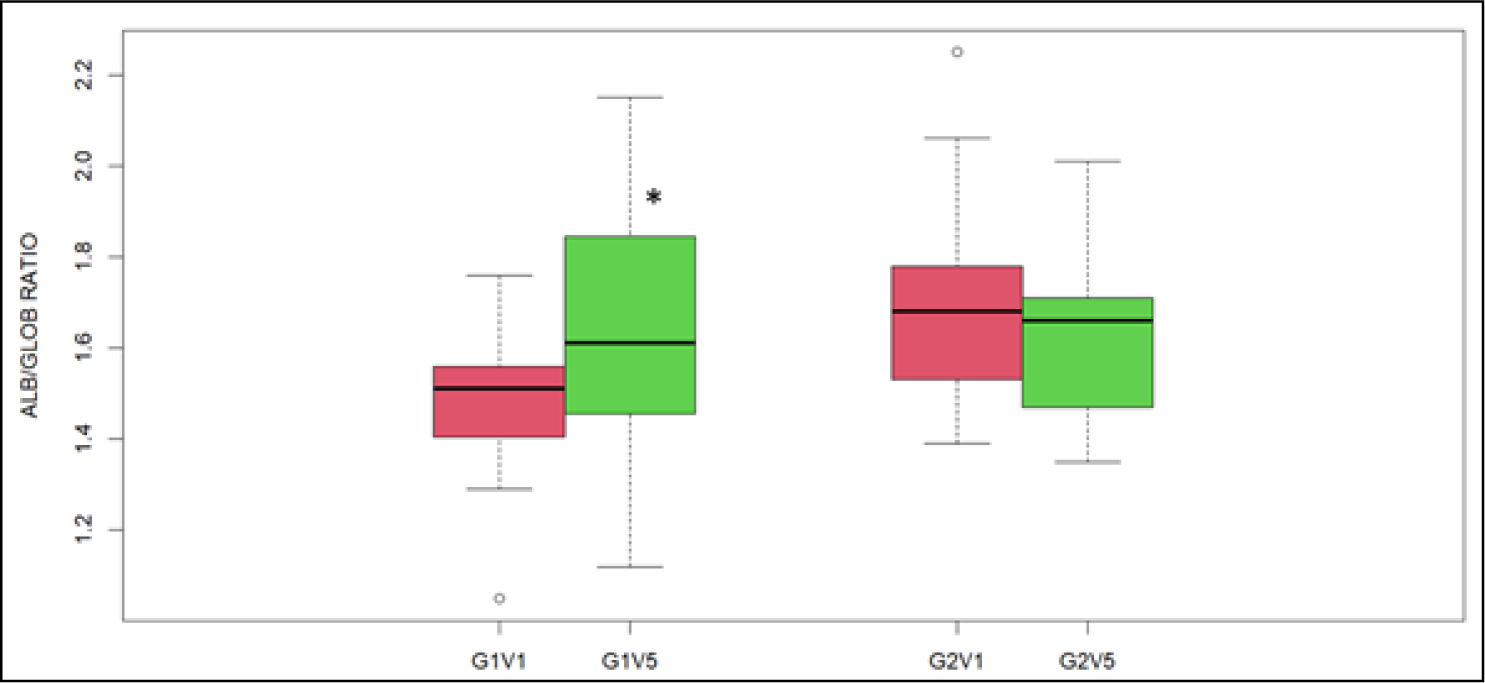
Effect of Lutein and Zeaxanthin supplementation on Albumin Globulin ratio levels

**Table 25:** Albumin to Globulin ratio during Visit-1 in group-1 (G1, treatment group) and group-2 (G2, placebo group)

| Albumin-Globulin ratio | G1 (N=16) | G2 (N=14) | Overall (N=30) |
| --- | --- | --- | --- |
| Mean (SD) | 1.49 (0.174) | 1.70 (0.229) | 1.59 (0.225) |
| Median (Min, Max) | 1.51 (1.05, 1.76) | 1.68 (1.39, 2.25) | 1.54 (1.05, 2.25) |
| Missing | 1 (6.3%) | 0 (0%) | 1 (3.3%) |

**Table 26:** Albumin to Globulin ratio during Visit-5 in group-1 (G1, treatment group) and group-2 (G2, placebo group)

| Albumin-Globulin ratio | G1 (N=16) | G2 (N=14) | Overall (N=30) |
| --- | --- | --- | --- |
| Mean (SD) | 1.62 (0.260) | 1.63 (0.206) | 1.62 (0.233) |
| Median (Min, Max) | 1.61 (1.12, 2.15) | 1.66 (1.35, 2.01) | 1.65 (1.12, 2.15) |
| Missing | 0 (0%) | 1 (7.1%) | 1 (3.3%) |

G1V1 (Alb/Glob ratio measured in group-1 at visit-1); G1V5 (Alb/Glob ratio measured in group-1 at visit-5); G2V1 (Alb/Glob ratio measured in group-2 at visit-1); G2V5 (Alb/Glob ratio measured in group-2 at visit-5). The central line in the box plots represents the median values, and the rectangular box shows the interquartile range (IQR), which is the middle 50% of the data. The whiskers extend to the minimum and maximum values of the data, except for the outliers that fall beyond the whiskers. Statistical significance was calculated using a paired, two-sided t-test. * indicates p<0.05

### 3.4 Total Bilirubin

The total bilirubin levels for G1 and G2 were 0.609 and 0.547 mg/dL, respectively, during visit 1. The corresponding values during visit 5 were 0.633 and 0.662 for G1 and G2, respectively. The comparisons are represented in Tables 27 and 28 and graphically in Figure 11.

**Figure 11:**
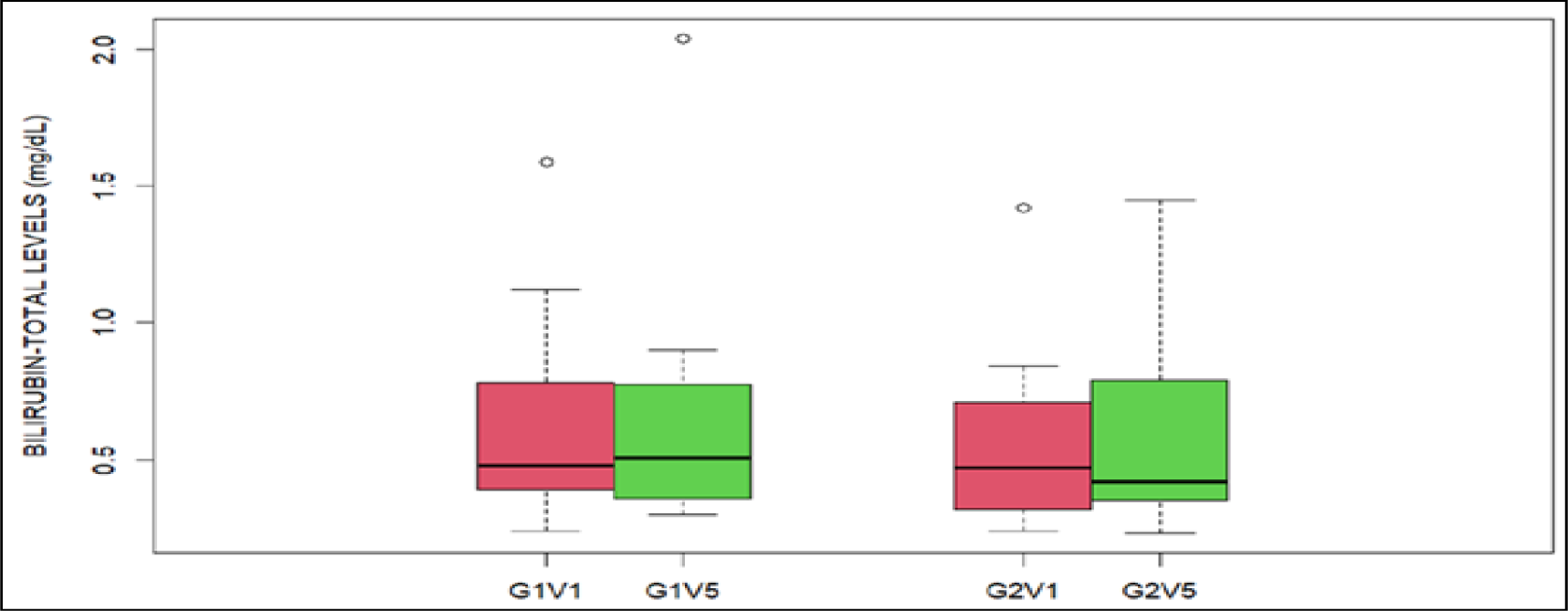
Effect of Lutein and Zeaxanthin supplementation on Total-Bilirubin levels

**Table 27:** Bilirubin (total) levels (mg/dL) during Visit-1 in group-1 (G1, treatment group) and group-2 (G2, placebo group)

| Total bilirubin | G1 (N=16) | G2 (N=14) | Overall (N=30) |
| --- | --- | --- | --- |
| Mean (SD) | 0.609 (0.359) | 0.547 (0.315) | 0.580 (0.335) |
| Median (Min, Max) | 0.480 (0.240, 1.59) | 0.470 (0.240, 1.42) | 0.470 (0.240, 1.59) |

**Table 28:** Bilirubin (total) levels (mg/dL) during Visit-5 in group-1 (G1, treatment group) and group-2 (G2, placebo group)

| Total bilirubin | G1 (N=16) | G2 (N=14) | Overall (N=30) |
| --- | --- | --- | --- |
| Mean (SD) | 0.633 (0.426) | 0.662 (0.427) | 0.646 (0.419) |
| Median (Min, Max) | 0.505 (0.300, 2.04) | 0.420 (0.230, 1.45) | 0.500 (0.230, 2.04) |
| Missing | 0 (0%) | 1 (7.1%) | 1 (3.3%) |

G1V1 (bilirubin-total levels measured in group-1 at visit-1); G1V5 (bilirubin-total levels measured in group-1 at visit-5); G2V1 (bilirubin-total levels measured in group-2 at visit-1); G2V5 (bilirubin-total levels measured in group-2 at visit-5). The central line in the box plots represents the median values, and the rectangular box shows the interquartile range (IQR), which is the middle 50% of the data. The whiskers extend to the minimum and maximum values of the data, except for the outliers that fall beyond the whiskers. Statistical significance was calculated using a paired, two-sided t-test. No significance was observed for corresponding comparisons.

### 3.5 Direct bilirubin

Tables 29 and 30 showed the total bilirubin levels for G1 and G2 were 0.183 and 0.176 mg/dL, respectively, during visit 1. The corresponding values during visit 5 were 0.217 and 0.219 mg/dL for G1 and G2, respectively. The comparisons are represented graphically in Figure 12.

**Figure 12:**
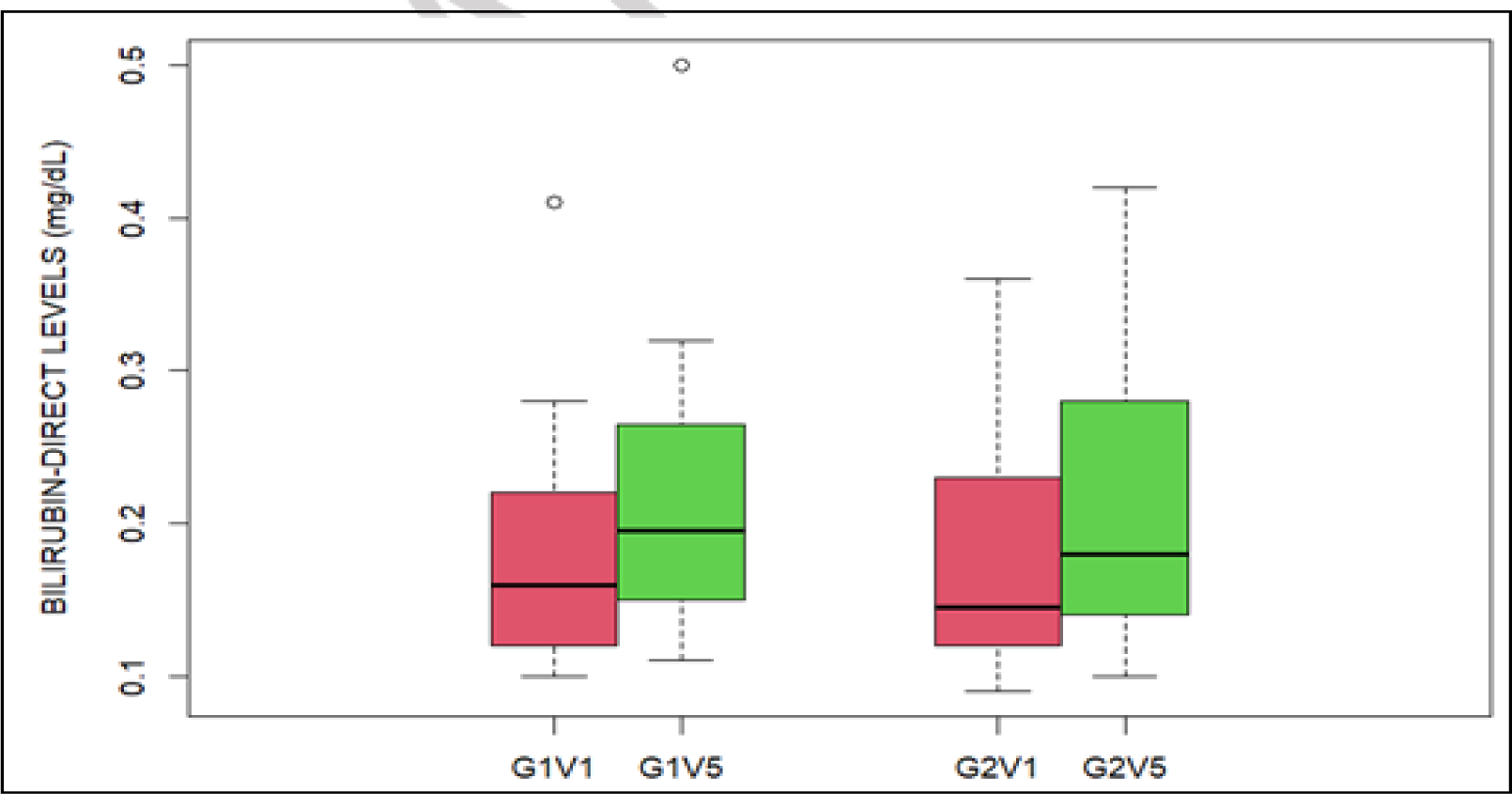
Effect of Lutein and Zeaxanthin supplementation on Bilirubin direct levels

**Table 29:** Bilirubin (direct) levels (mg/dL) during Visit-1 in group-1 (G1, treatment group) and group-2 (G2, placebo group)

| Bilirubin direct | G1 (N=16) | G2 (N=14) | Overall (N=30) |
| --- | --- | --- | --- |
| Mean (SD) | 0.183 (0.0814) | 0.176 (0.0803) | 0.180 (0.0795) |
| Median (Min, Max) | 0.160 (0.100, 0.410) | 0.145 (0.0900, 0.360) | 0.155 (0.0900, 0.410) |

**Table 30:**
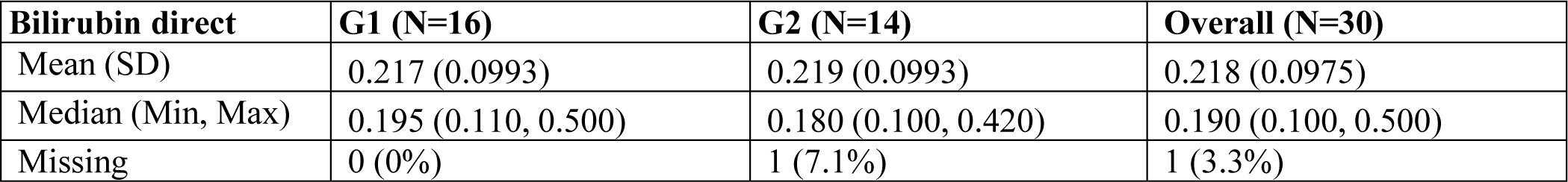
Bilirubin (direct) levels (mg/dL) during Visit-5 in group-1 (G1, treatment group) and group-2 (G2, placebo group)

| Bilirubin direct | G1 (N=16) | G2 (N=14) | Overall (N=30) |
| --- | --- | --- | --- |
| Mean (SD) | 0.217 (0.0993) | 0.219 (0.0993) | 0.218 (0.0975) |
| Median (Min, Max) | 0.195 (0.110, 0.500) | 0.180 (0.100, 0.420) | 0.190 (0.100, 0.500) |
| Missing | 0 (0%) | 1 (7.1%) | 1 (3.3%) |

G1V1 (bilirubin-direct levels measured in group-1 at visit-1); G1V5 (bilirubin-direct levels measured in group-1 at visit-5); G2V1 (bilirubin-direct levels measured in group-2 at visit-1); G2V5 (bilirubin-direct levels measured in group-2 at visit-5). The central line in the box plots represents the median values, and the rectangular box shows the interquartile range (IQR), which is the middle 50% of the data. The whiskers extend to the minimum and maximum values of the data, except for the outliers that fall beyond the whiskers.

### 3.6 Indirect bilirubin

Tables 31 and 32 show the indirect bilirubin levels for G1 and G2 during visits 1 and 5.

**Table 31:**
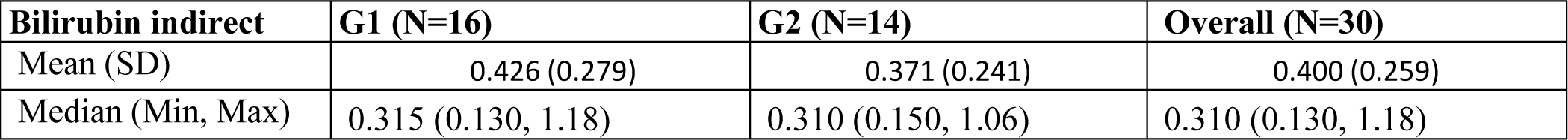
Bilirubin(indirect) levels (mg/dL) during Visit-1 in group-1 (G1, treatment group) and group-2 (G2, placebo group)

**Table 32:** Bilirubin (indirect) levels (mg/dL) during Visit-5 in group-1 (G1, treatment group) and group-2 (G2, placebo group)

| **Bilirubin indirect** | **G1 (N=16)** | **G2 (N=14)** | **Overall (N=30)** |
| --- | --- | --- | --- |
| Mean (SD) | 0.416 (0.333) | 0.443 (0.333) | 0.428 (0.327) |
| Median (Min, Max) | 0.325 (0.190, 1.54) | 0.280 (0.100, 1.06) | 0.310 (0.100, 1.54) |
| Missing | 0 (0%) | 1 (7.1%) | 1 (3.3%) |

### 3.7 SGPT and SGOT

The mean values of SGPT in different groups of participants during visits 1 and 2 are presented in tables 33 and 34 and Figure 13. The mean values of SGOT in different groups of participants during visits 1 and 2 are presented in tables 35 and 36 and Figure 14.

**Figure 13:**
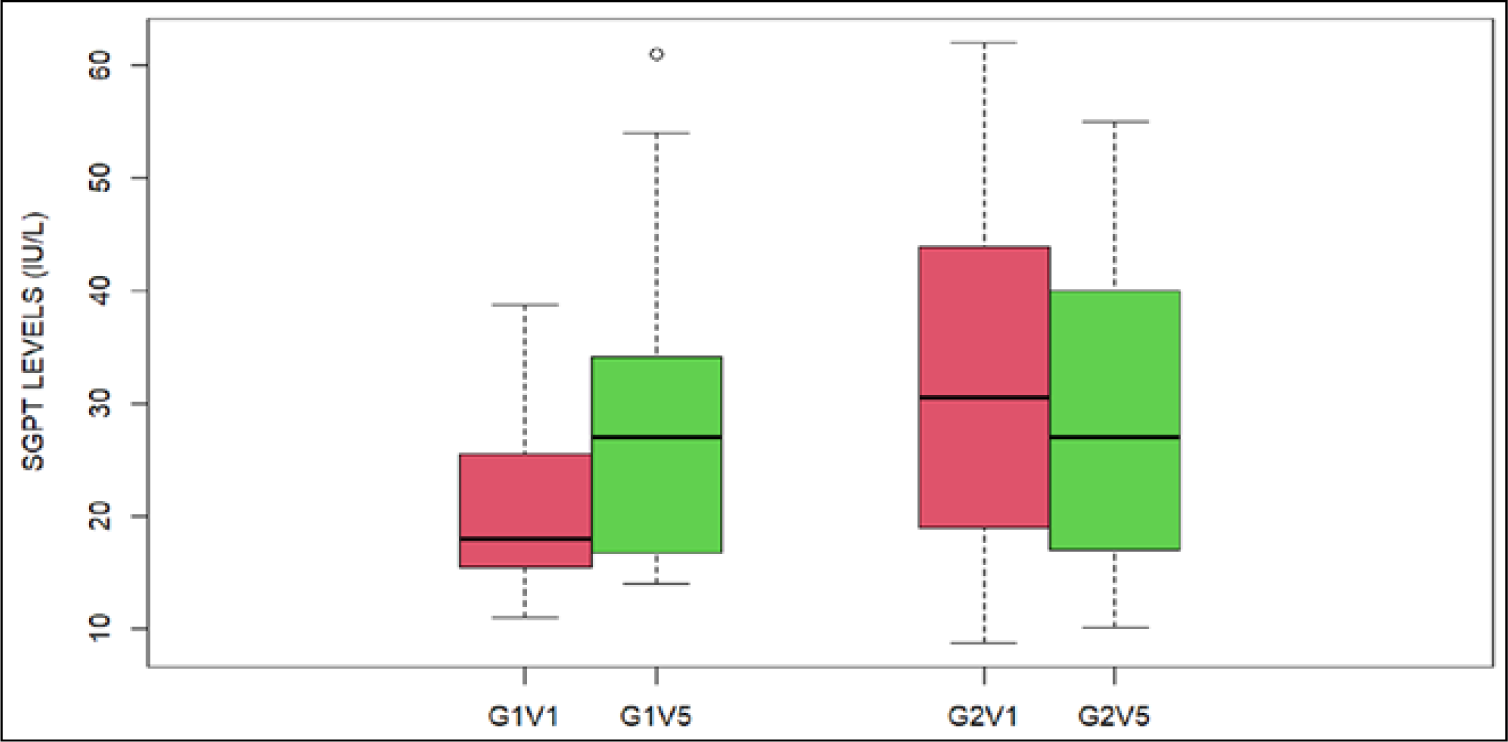
Effect of Lutein and Zeaxanthin supplementation on SGPT levels

**Figure 14:**
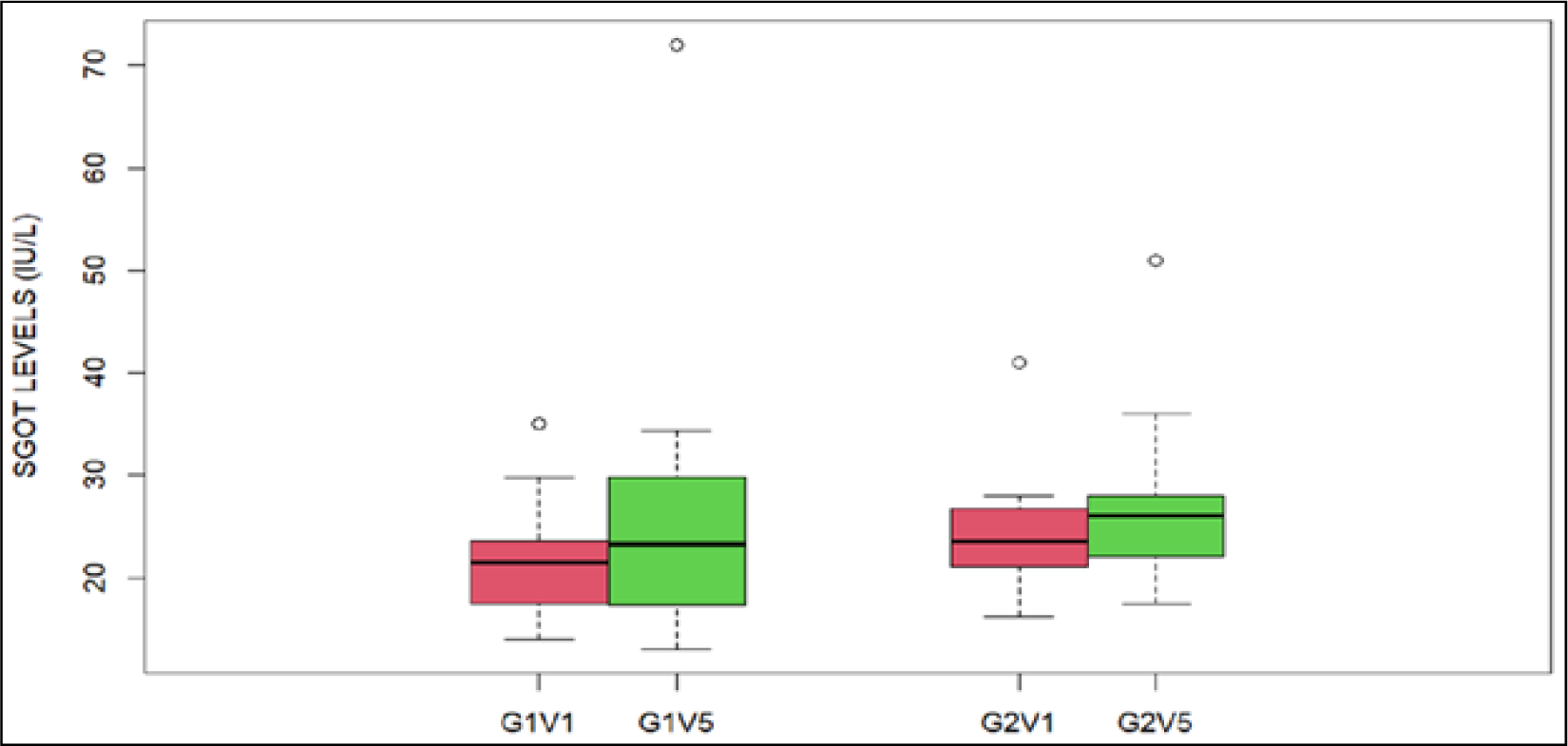
Effect of Lutein and Zeaxanthin supplementation on SGOT levels

**Table 33:** SGPT levels (IU/L) during Visit-1 in group-1 (G1, treatment group) and group-2 (G2, placebo group)

| **SGPT** | **G1 (N=16)** | **G2 (N=14)** | **Overall (N=30)** |
| --- | --- | --- | --- |
| Mean (SD) | 20.4 (7.36) | 31.4 (15.2) | 25.5 (12.8) |
| Median (Min, Max) | 18.0 (11.0, 38.8) | 30.5 (8.70, 62.0) | 23.0 (8.70, 62.0) |

**Table 34:** SGPT levels (IU/L) during Visit-5 in group-1 (G1, treatment group) and group-2 (G2, placebo group)

| **SGPT** | **G1 (N=16)** | **G2 (N=14)** | **Overall (N=30)** |
| --- | --- | --- | --- |
| Mean (SD) | 28.2 (14.2) | 30.2 (13.7) | 29.1 (13.7) |
| Median (Min, Max) | 27.0 [14.0, 61.0] | 27.0 [10.1, 55.0] | 27.0 [10.1, 61.0] |
| Missing | 0 (0%) | 1 (7.1%) | 1 (3.3%) |

**Table 35:** SGOT levels (IU/L) during Visit-1 in group-1 (G1, treatment group) and group-2 (G2, placebo group)

| SGOT | G1 (N=16) | G2 (N=14) | Overall (N=30) |
| --- | --- | --- | --- |
| Mean (SD) | 21.5 (5.79) | 24.3 (5.88) | 22.8 (5.90) |
| Median (Min, Max) | 21.5 (14.0, 35.0) | 23.5 (16.2, 41.0) | 22.0 (14.0, 41.0) |

**Table 36:** SGOT levels (IU/L) during Visit-5 in group-1 (G1, treatment group) and group-2 (G2, placebo group)

| SGOT | G1 (N=16) | G2 (N=14) | Overall (N=30) |
| --- | --- | --- | --- |
| Mean (SD) | 26.1 (13.9) | 27.0 (8.70) | 26.5 (11.6) |
| Median (Min, Max) | 23.3 (13.0, 72.0) | 26.0 (17.4, 51.0) | 25.0 (13.0, 72.0) |
| Missing | 0 (0%) | 1 (7.1%) | 1 (3.3%) |

G1V1 (SGPT levels measured in group-1 at visit-1); G1V5 (SGPT levels measured in group-1 at visit-5); G2V1 (SGPT levels measured in group-2 at visit-1); G2V5 (SGPT levels measured in group-2 at visit-5). The central line in the box plots represents the median values, and the rectangular box shows the interquartile range (IQR), which is the middle 50% of the data. The whiskers extend to the minimum and maximum values of the data, except for the outliers that fall beyond the whiskers. Statistical significance was calculated using a paired, two-sided t-test. No significance was observed for corresponding comparisons.

G1V1 (SGOT levels measured in group-1 at visit-1); G1V5 (SGOT levels measured in group-1 at visit-5); G2V1 (SGOT levels measured in group-2 at visit-1); G2V5 (SGOT levels measured in group-2 at visit-5). The central line in the box plots represents the median values, and the rectangular box shows the interquartile range (IQR), which is the middle 50% of the data. The whiskers extend to the minimum and maximum values of the data, except for the outliers that fall beyond the whiskers. Statistical significance was calculated using a paired, two-sided t-test. No significance was observed for corresponding comparisons.

### 3.8 Alkaline phosphatase (ALP)

In Tables 37 and 38 and Figure 15 below, the mean ALP levels increased from 77.9 to 87.2 IU/L in G-1 between visits 1 and 5. Whereas in G-2, the mean ALP levels were 70.2 and 68.4 IU/L for visits 1 and 5, respectively. These changes were statistically significant as estimated by a paired, two-tailed t-test.

**Figure 15:**
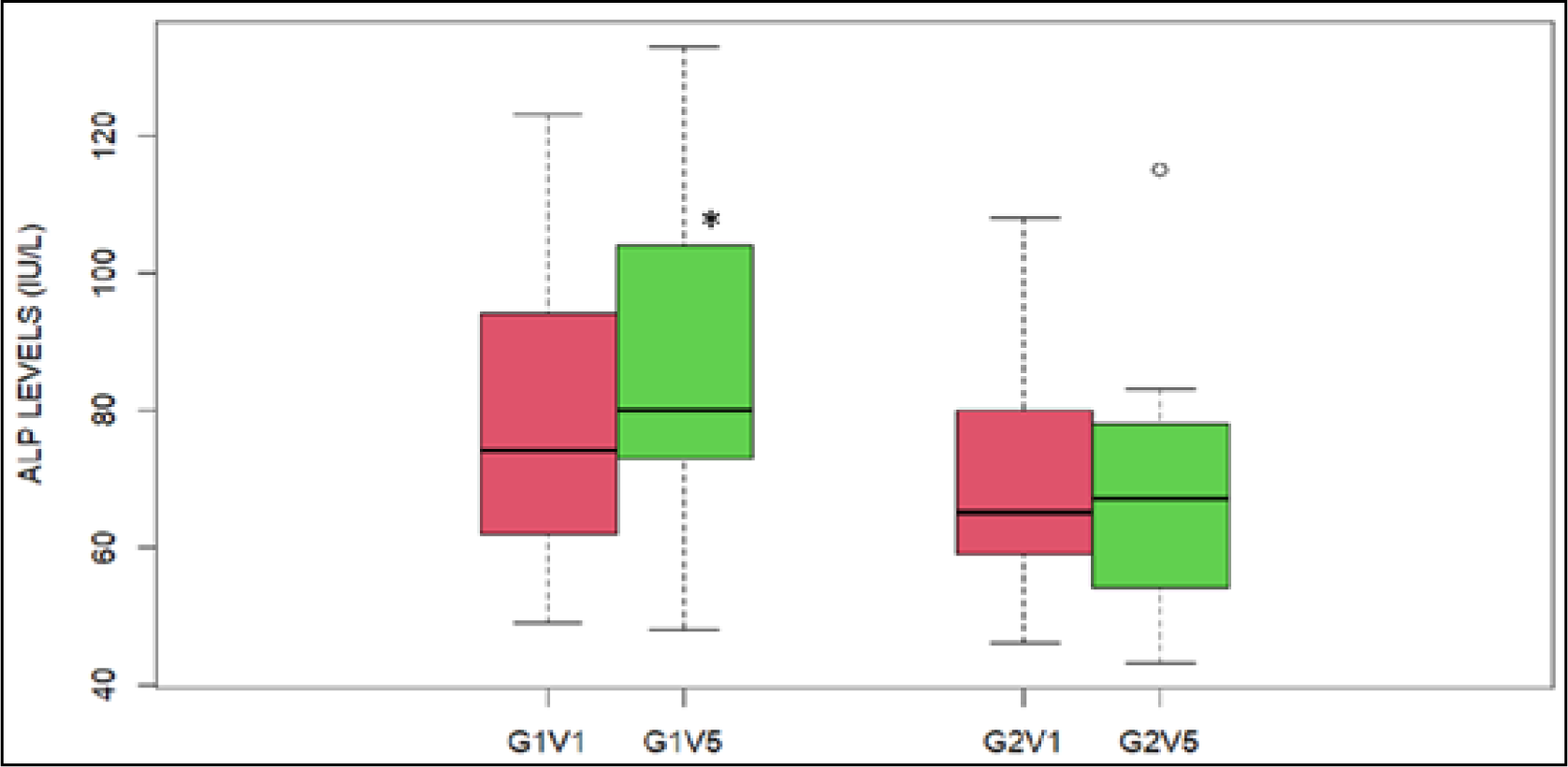
Effect of Lutein and Zeaxanthin supplementation on ALP levels

**Table 37:** ALP levels (IU/L) during Visit-1 in group-1 (G1, treatment group) and group-2 (G2, placebo group)

| ALP | G1 (N=16) | G2 (N=14) | Overall (N=30) |
| --- | --- | --- | --- |
| Mean (SD) | 77.9 (21.1) | 70.2 (16.3) | 74.2 (19.0) |
| Median (Min, Max) | 74.0 (49.0, 123) | 65.0 (46.0, 108) | 67.0 (46.0, 123) |
| Missing | 1 (6.3%) | 0 (0%) | 1 (3.3%) |

**Table 38:** ALP levels (IU/L) during Visit-5 in group-1 (G1, treatment group) and group-2 (G2, placebo group)

| ALP | G1 (N=16) | G2 (N=14) | Overall (N=30) |
| --- | --- | --- | --- |
| Mean (SD) | 87.2 (25.7) | 68.4 (18.7) | 78.8 (24.4) |
| Median (Min, Max) | 80.0 (48.0, 133) | 67.0 (43.0, 115) | 75.0 (43.0, 133) |
| Missing | 0 (0%) | 1 (7.1%) | 1 (3.3%) |

G1V1 (ALP levels measured in group-1 at visit-1); G1V5 (ALP levels measured in group-1 at visit-5); G2V1 (ALP levels measured in group-2 at visit-1); G2V5 (ALP levels measured in group-2 at visit-5). The central line in the box plots represents the median values, and the rectangular box shows the interquartile range (IQR), which is the middle 50% of the data. The whiskers extend to the minimum and maximum values of the data, except for the outliers that fall beyond the whiskers. Statistical significance was calculated using a paired, two-sided t-test. P<0.05.

### 3.9 Gamma Glutamyl Transpeptidase (GGTP)

The mean GGTP levels for G1 were 25.9 and 29.6 IU/L for visits 1 and 5, respectively. Similarly, for G2, the GGTP levels were 33.3 and 34.8 IU/L for visits 1 and 5, respectively. The differences were not statistically significant (Tables 39 and 40 and Figure 16).

**Figure 16:**
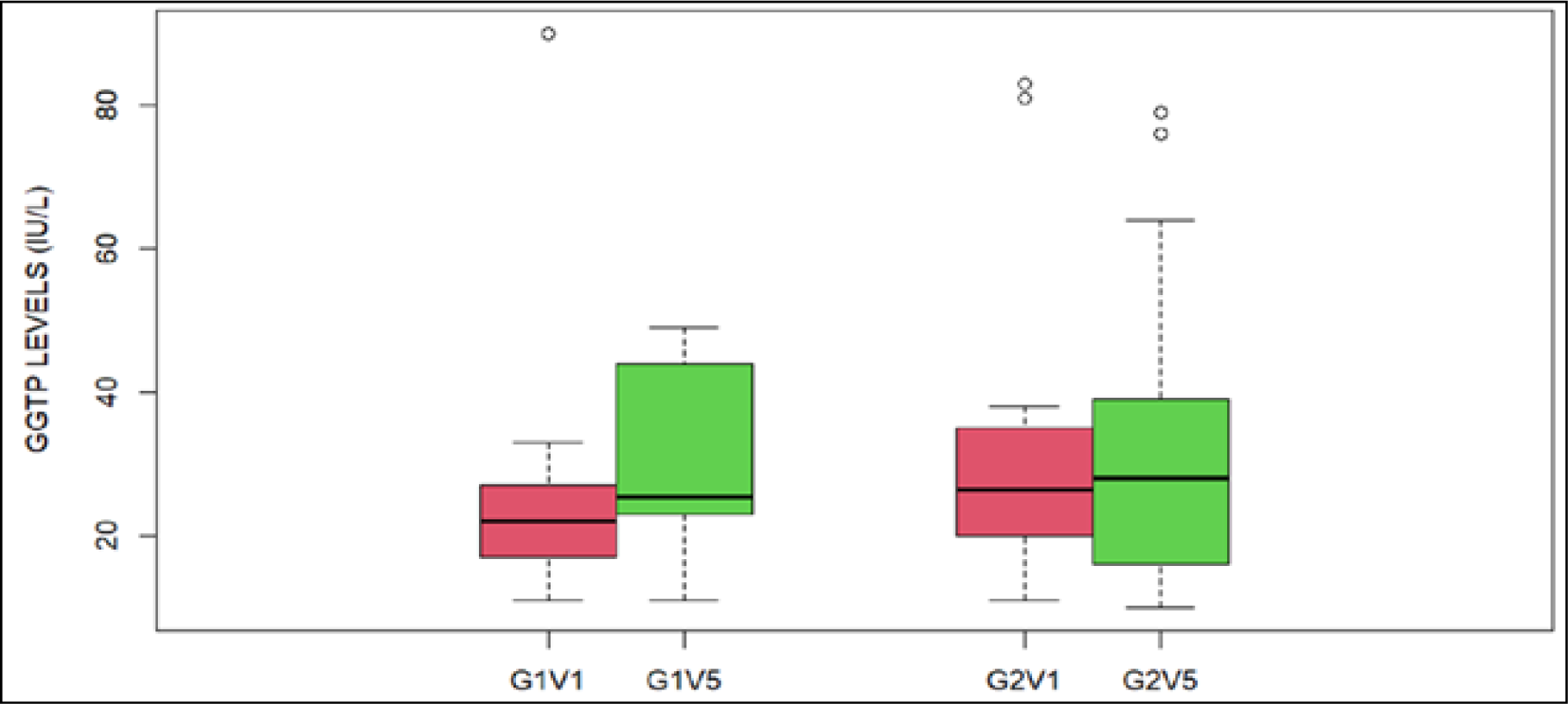
Effect of Lutein and Zeaxanthin supplementation on GGTP levels

**Table 39:** GGTP levels (IU/L) during Visit-1 in group-1 (G1, treatment group) and group-2 (G2, placebo group)

| GGTP | G1 (N=16) | G2 (N=14) | Overall (N=30) |
| --- | --- | --- | --- |
| Mean (SD) | 25.9 (18.8) | 33.3 (22.0) | 29.5 (20.4) |
| Median (Min, Max) | 22.0 (11.0, 90.0) | 26.5 (11.0, 83.0) | 25.0 (11.0, 90.0) |
| Missing | 1 (6.3%) | 0 (0%) | 1 (3.3%) |

**Table 40:** GGTP levels (IU/L) during Visit-5 in group-1 (G1, treatment group) and group-2 (G2, placebo group)

| GGTP | G1 (N=16) | G2 (N=14) | Overall (N=30) |
| --- | --- | --- | --- |
| Mean (SD) | 29.6 (12.5) | 34.8 (23.5) | 31.9 (18.1) |
| Median (Min, Max) | 25.5 (11.0, 49.0) | 28.0 (10.0, 79.0) | 27.0 (10.0, 79.0) |
| Missing | 0 (0%) | 1 (7.1%) | 1 (3.3%) |

G1V1 (GGTP levels measured in group-1 at visit-1); G1V5 (GGTP levels measured in group-1 at visit-5); G2V1 (GGTP levels measured in group-2 at visit-1); G2V5 (GGTP levels measured in group-2 at visit-5). The central line in the box plots represents the median values, and the rectangular box shows the interquartile range (IQR), which is the middle 50% of the data. The whiskers extend to the minimum and maximum values of the data, except for the outliers that fall beyond the whiskers. Statistical significance was calculated using a paired, two-sided t-test, and there were no significant differences between either of the groups for visits 1 and 5. And here the Table 41 were showing the criteria for classifying the outcome of study in liver injury.

**Table 41:** The criteria for classifying the outcome of study in liver injury.

| Type of injury | Enzymatic profile |
| --- | --- |
| Hepatocellular | ALT > 2ULN<br>Serum ALT/Serum Alk.<br>Phos $\geq 5^*$ |
| Cholestatic | Alk Phos $\geq 2$ULN<br>Serum ALT/Serum Alk<br>Phos $\leq 2^*$ |
| Mixed | ALT > 2 ULN<br>Serum ALT/Serum Alk<br>Phos between 2 and 5* |

Based on the above results LFT to represent hepatocellular injury in the ALT levels should be twice more than the upper limit of normal (ULN) and similarly there are specific criteria for identifying cholestatic and mixed types of injury. In our studies, most of the individuals either in the group-1 or group-2 did not show increase corresponding to more than twice of ULN for these parameters. In addition, for neither of the groups the LFT parameters showed any statistically significant increase.

## DISCUSSION

Numerous research evaluated the impact of lutein on normal and diabetic rats. It was discovered that diabetic rats with dose-dependent extract ingestion had lower blood glucose levels than their healthy equivalents. According to the Hosseini et al. study, when compared to the values at the start of the trial and those of the control group, the levels of FBS, HbA1c, total cholesterol, and triglycerides dramatically decreased in the diabetic patients receiving lutein treatment (38). The American Diabetes Association advises using HbA1c with a cut-point 6.5% for diagnosing diabetes as an alternative to fasting plasma glucose since it offers a reliable marker of chronic glycemia and corresponds well with the risk of long-term diabetes complications (56). Additionally, HbA1c is an excellent predictor of lipid profile, offering the additional benefit of recognizing cardiovascular risk in diabetic patients (57).

So, when the Lutein and zeaxanthin were compared to placebo, they reduced the HbA1c levels. The analysis of HbA1c levels, estimated average glucose levels (eAG), and random glucose levels showed good results in newly diagnosed diabetic patients. These results suggest that Lutein and zeaxanthin can be a beneficial supplement for diabetic patients with type 2 diabetes. Hypertension, aging, hemodynamic dysregulation, proteinuria, and a high consumption of dietary protein are some of the conditions that may accelerate kidney damage. A significant risk factor for the advancement of CKD, cardiovascular disease, and all-cause mortality has been found to be proteinuria, which is unrelated to kidney disease. Effective proteinuria management is important as a key therapy response indicator in a number of renal disorders. Kidney function tests are utilized in this clinical research to evaluate the safety profile of the supplement and any potential kidney side effects. The tests aid in figuring out whether the supplement is having an impact on kidney function indicators including creatinine, blood urea nitrogen, and serum urea. A common laboratory test to determine the level of urea nitrogen in the blood is the blood urea nitrogen (BUN) test. The waste product urea is expelled by the kidneys and is created when proteins are broken down in the liver. The study results indicate that treatment with lutein did not increase blood urea nitrogen. Urea is primarily eliminated via the kidneys, with the remaining portion excreted through the gastrointestinal tract. Elevated levels of urea in the blood may indicate renal impairment or other conditions such as dehydration, high protein diets, and upper GI bleeding. In contrast, decreased levels of urea may indicate low-protein diets, starvation, or severe liver disease. The serum urea level is a commonly used laboratory test to assess kidney function and monitor kidney disease. Serum creatinine level is another parameter commonly measured in blood tests to assess kidney function, as the kidneys are responsible for filtering and excreting creatinine from the body. Elevated serum creatinine levels can indicate decreased kidney function or damage to the kidneys, while low levels may indicate decreased muscle mass or other medical conditions. However, the interpretation of serum creatinine levels should be done in conjunction with other clinical information, such as the patient’s age, sex, weight, and medical history. The serum creatinine levels did not show any significant variation in either of the groups during visit 5 in comparison to visit 1, according to the findings of this study. These parameters can indicate whether the supplement is causing damage to the kidneys or affecting their ability to function correctly. Therefore, it is essential to monitor kidney function parameters regularly during clinical trials.

In India, a significant number of laboratories rely on RI (reference intervals) derived from literature that primarily focuses on Western populations. However, the applicability of these RI is questionable due to disparities stemming from differences in diet, lifestyle, environmental conditions, and ethnicity-related variations. Keeping this in mind, a recent study was conducted on 2,021 healthy individuals in India to provide reference intervals (RI) for certain liver-specific biochemical parameters (58). The study measured several blood parameters and reported reference ranges for several liver-specific parameters. According to this study, bilirubin had a wide variation in the observed RI (0.30–1.30) compared to the reported RI (0– 1.2). Similarly, SGOT showed a wide variation (13-52.80) compared to the reported RI (0–40). SGPT and ALP also had wide variations in the observed RI (10–68 for SGPT and 107–361.80 for ALP) compared to their reported standard RI (0–50 for SGPT and 110–310 for ALP). However, GGT, a marker for biliary disease and alcoholism, remained within the reported standard RI (0–50, 5.00–50.60). The total protein test is useful in identifying various health conditions, such as liver disease, kidney disease, and malnutrition resulting from inadequate nutrient intake. Abnormally low levels of total protein may indicate a problem with protein digestion or absorption or a liver or kidney disorder. In both groups, the total protein levels did not decrease, indicating that the treatment did not have any negative effects. In fact, in both conditions, the total protein levels increased, and this increase was statistically significant. However, the interpretation of an increase in total protein in a physiological context is complex. From the results, we can only infer that the total protein levels did not reduce during the treatment. Serum albumin is a protein that is produced by the liver and is found in the blood. It plays an essential role in maintaining the osmotic pressure of the blood and helps to transport various substances, including hormones, drugs, and fatty acids. Low levels of serum albumin can indicate liver or kidney disease, malnutrition, or inflammation. Monitoring serum albumin levels during a clinical trial is critical in order to evaluate the effects of the intervention on liver function. There are several drugs that can lower serum albumin levels, either by reducing albumin production or increasing its breakdown. In both groups, the albumin levels did not decrease, indicating that the treatment did not have any negative effects. In fact, in G1, albumin levels increased, and this increase was statistically significant. However, the interpretation of an increase in total protein in a physiological context is complex. From the results, we can only infer that the total protein levels did not reduce during the treatment. In either G1 or G2, during the clinical trial, the serum globulin levels did not drop below 2 mg/dL, indicating that the treatment did not have negative effects. Albumin is the most abundant protein in the blood and plays a crucial role in maintaining oncotic pressure, transporting various substances (such as hormones and drugs), and regulating fluid balance. Globulins, on the other hand, are a diverse group of proteins involved in immune function, blood clotting, and transporting other substances. The A/G ratio is used as a marker to evaluate liver and kidney function, nutritional status, and certain medical conditions. In liver disease, impaired albumin production leads to a decreased A/G ratio. Kidney damage affects protein filtration, altering the A/G ratio. Inflammation and infection can raise certain globulin levels, increasing the A/G ratio. Total bilirubin, indirect bilirubin, and direct bilirubin are the essential markers of liver health and function, as elevated levels of total bilirubin can indicate liver damage or dysfunction, as well as other medical conditions affecting the liver.

Both SGPT (Serum Glutamate Pyruvate Transaminase, or ALT) and SGOT (Serum Glutamic Oxaloacetic Transaminase, or AST) are liver enzymes commonly used in liver function tests. While both enzymes provide valuable information about liver health, their interpretations and clinical significance differ. SGPT (ALT) is primarily found in the liver, and elevated levels typically indicate liver damage or injury. It is considered more specific to liver function, making it a reliable marker for hepatocellular damage such as viral hepatitis, liver cirrhosis, or drug-induced liver injury. SGOT (AST), on the other hand, is present not only in the liver but also in other organs like the heart, skeletal muscles, and kidneys. Therefore, elevated SGOT levels may not solely reflect liver-specific issues. Increased SGOT levels can be associated with liver damage but can also be caused by conditions affecting other organs or tissues.

ALP (alkaline phosphatase) is a hydrolase enzyme that is widely distributed in various tissues, including the liver, bone, intestine, and placenta. In the context of liver function tests during clinical safety validation, ALP is primarily used as a marker for hepatobiliary system evaluation. Elevated ALP levels can indicate cholestasis, obstructive liver diseases, or hepatic cell injury. It is particularly useful in distinguishing between hepatocellular and cholestatic liver disorders. ALP measurement aids in assessing liver health, monitoring disease progression, and determining the potential hepatotoxic effects of drugs. In clinical practice, ALP serves as a valuable biochemical parameter to enhance the diagnostic accuracy and safety assessment of liver function. GGTP is a microsomal enzyme found in various cells, including hepatocytes, biliary epithelial cells, renal tubules, the pancreas, and the intestine. It plays a role in transporting peptides across the cell membrane and is involved in glutathione metabolism. Although GGTP is present in renal tissue, its activity in the serum is primarily associated with the hepatobiliary system. Acute viral hepatitis and alcoholism can significantly increase the levels of GGTP. Additionally, GGTP can serve as an early marker of oxidative stress.

Measurements of bone mineral density and clinical risk factors are now utilized to determine who is at risk for osteoporosis. ALP is an enzyme that is made in the liver, bones, intestines, and kidneys and gets into the bloodstream. Total serum ALP levels as a bone-forming biomarker have been shown in studies to be able to predict how well medication therapy for osteoporosis is working (59, 60). Today, there have been some documented clinical trials on how people with osteoporosis respond to herbal treatment. A recent Cochrane systematic review showed that most included studies (85 of 108) had small sample sizes, ranging from 20 to 120, and that the overall quality of the evidence was generally low. The use of herbal medicine among people with osteoporosis is therefore not supported by clear evidence, and larger sample sizes and more thoroughly planned research are needed. It should be mentioned that alkaline phosphatase (ALP) is a crucial enzyme that is essential for the development of hard tissue, especially mineralized tissue, as well as for liver function. According to research, extracellular pyrophosphate, which is an inhibitor of mineral formation, and inorganic phosphate local rates are both increased by ALP, which promotes mineralization (61). Total alkaline phosphatase levels are actually shown to be a measure of increased osteoblastic activity and bone conversion status (62). According to a study by Takeda et al., four weeks of lutein treatment significantly increased the mass of the femoral bones, particularly the cortical bone. This improvement was quantified by dual X-ray absorptiometry and micro computed tomography (CT) assessments of bone mineral density (63). Male hip fracture risk has been demonstrated to be negatively linked with dietary intake of the carotenoids α-carotene, β-carotene, and lutein (64). Similar findings were made by Zhang et al (65) who discovered that higher serum carotenoid levels were associated with greater bone mineral density in both men and women. Lutein, which is well known for its antioxidant abilities, contributes to bone remodelling by acting as an antioxidant. Our research suggests that supplementing with Lutein and zeaxanthin may have a beneficial effect on bone formation. However, more research is need to fully understand this occurrence.

## CONCLUSION

In this study, the effects of Lutein and zeaxanthin on blood sugar levels, kidney, liver, and bone health are described in depth. Five weeks of lutein supplementation improved the levels of HbA1c. When examining the safety and effectiveness of supplements in clinical studies, kidney function tests are essential. This is so that waste and extra fluid can be removed from the body, a task that the kidneys are extremely important for performing. The build-up of toxins in the body due to impaired renal function can have negative impacts on health. It is possible that lutein supplementation is not harmful to renal function, according to parameters such as blood urea nitrogen, serum urea levels, and creatinine levels. Our clinical investigation revealed that the Lutein and zeaxanthin had a good impact on these kidney function markers and was safe. The liver function tests suggest that using lutein supplements may not pose any hazards for the liver. The SGOT and SGPT are the key factors in identifying the levels of liver damage. Alkaline phosphatase levels increased when lutein was administered. However, the level of the ALP increase was not sufficiently significant to be categorized as a liver damage case. Lutein supplementation is known to be beneficial for bone growth. Considering the role of ALP (Alkaline Phosphatase) in bone mineralisation, the small change in ALP in this study indicates that Lutein may have effect in improving the bone health and yet at the same time the increase in the values of ALP was not statistically significant proving the safety aspects of lutein.

However, it is hoped that through this meticulous validation, the findings of this randomized controlled trial would demonstrate the effectiveness and safety of Lutein and zeaxanthin for bone health and serve as a solid justification for conducting additional research for better activity.

## Data Availability

All data produced in the present work are contained in the manuscript

## Notes

### Competing Interest Statement

The authors have declared no competing interest.

### Clinical Trial

CTRI/2022/06/043208

### Funding Statement

This study did not receive any funding

### Author Declarations

The ethical clearance was obtained at Mangala Institutional Ethics Committee vide letter Ref. No. MIEC/V6.1/015

